# Wide-angled proteomics in giant cell arteritis reveals complement pathway activation

**DOI:** 10.64898/2026.10.14.26365544

**Authors:** Benjamin J. Whittle, Martina Williams, Hannah Lomax-Browne, Louise Sorensen, Robert T. Maughan, Matthew Porteous, Aruna Chakrabarty, Raashid A. Luqmani, Matthew C. Pickering, Ann W. Morgan, James E. Peters

**Affiliations:** Department of Immunology and Inflammation, Imperial College London, United Kingdom; School of Medicine, University of Leeds and NIHR Leeds Biomedical Research Centre, Leeds Teaching Hospitals NHS Trust, Leeds, United Kingdom; Nuffield Department of Orthopaedics, Rheumatology and Musculoskeletal Science, University of Oxford, Oxford, United Kingdom

## Abstract

Giant cell arteritis (GCA) is an immune-mediated vasculitis that can cause serious morbidity including sight loss, stroke and aortic aneurysm. There is an unmet need for blood-based biomarkers and novel therapeutics. We measured plasma protein levels using the SomaScan v4.1 platform, comprising 7,273 protein measurements (SOMAmers) reflecting 6,596 unique protein targets. We profiled the proteome in 113 patients who presented acutely with possible GCA. 56 had biopsy-proven GCA and 57 did not have GCA (non-GCA). We found 144 SOMAmers increased and 151 reduced in GCA. Dysregulated biological pathways included IL6-JAK-STAT3 signalling, chemokines for both myeloid (e.g. CXCL2, CXCL3, CCL18, CCL23) and lymphoid cells (e.g. CXCL9, CXCL13), and the complement system (e.g. C9, CFHR5, C5a, Factor B). Enzyme-Linked Immunosorbent Assays (ELISAs) confirmed elevated plasma levels of C5a, the terminal complement complex, Factor B and CFHR5 in GCA. We developed a 26-protein signature that distinguished GCA from non-GCA (area-under-the curve, AUC, 0.93) and validated the diagnostic signature in external datasets. Our data implicate terminal complement pathway activation in GCA, raising the possibility of complement-directed therapies. We show proof-of-principle that a multiplex protein biomarker could be used as a diagnostic adjunct in suspected GCA.

## Introduction

Giant cell arteritis (GCA) is a vasculitis affecting large- and medium-sized arteries. Histopathologically, GCA is characterised by granulomatous arteritis, with a transmural inflammatory cell infiltrate, destruction of the elastic laminae, and intimal proliferation leading to luminal stenosis. Clinical manifestations result from systemic inflammation (e.g. weight loss, malaise, acute phase response), end-organ ischaemia (e.g. visual loss, stroke, limb claudication) or weakening of the arterial wall (aneurysm). Clinical classification systems subset GCA into cranial GCA (C-GCA) and large-vessel GCA (LV-GCA) based on the pattern of presenting symptoms and arterial territories involved. However, considerable overlap exists with estimates of large-vessel involvement of above 50% in C-GCA depending on how extensively a patient is imaged during the diagnostic work-up (1).

There are several challenges in the clinical management of GCA. First is the lack of GCA-specific blood-based diagnostic biomarkers. Prompt diagnosis and initiation of glucocorticoid treatment is critical to preventing irreversible visual loss (2). It is also important to avoid missing other serious alternative diagnoses and treating these inappropriately as GCA. Historically, the gold-standard diagnostic was a temporal artery biopsy (TAB), but increasingly this is being replaced by imaging (usually temporal artery and axillary ultrasound). Although ultrasound is non-invasive, it still presents logistical challenges and is not universally available. Moreover, the sensitivity of ultrasound rapidly decreases following glucocorticoid initiation. While inflammatory markers such as C-reactive protein (CRP) or Erythrocyte Sedimentation Rate (ESR) are sensitive, they are non-specific and are raised in infection and other inflammatory conditions (3–5). False positive diagnoses are associated with high healthcare costs (6). Thus, a novel blood-based biomarker that could be used to triage patients presenting with potential GCA would be a valuable clinical tool. A second challenge is that therapeutic progress in GCA lags behind that of other autoimmune rheumatic diseases (ARDs). In diseases such as rheumatoid arthritis, ANCA-associated vasculitis and lupus, the role of glucocorticoids has been increasingly relegated to acute control of disease, with glucocorticoid-free remission the goal. By contrast, in GCA glucocorticoid monotherapy is usually first-line therapy. Tocilizumab (7) and, more recently, upadacitinib (8) (a JAK inhibitor) have demonstrated efficacy as steroid-sparing agents in GCA, but the development of targeted therapies still trails behind other ARDs.

There have been rapid advances in proteomic technologies in recent years. Using a targeted proteomic platform, we previously measured 184 proteins in GCA (9) but coverage was limited to a small fraction of the circulating proteome and biased to inflammation and vascular-related proteins. Here, we expand to a more comprehensive assessment of the plasma proteome, measuring 7,273 targets corresponding to 6,596 unique proteins (**Figure 1**). Proteins upregulated in GCA implicate the IL6-JAK-STAT3 pathway and vascular remodelling consistent with known biology. We observed a complex chemokine signature spanning innate and adaptive immunity, with signals for neutrophil recruitment (CXCL2, CXCL3), monocytes/macrophages (CCL18, CCL23), IFN-γ-driven Th1 responses (CXCL9), and B-cell-associated lymphoid organisation (CXCL13). Strikingly, we identified strong upregulation of complement pathways and validated these proteomic findings using dedicated ELISAs, demonstrating evidence of terminal complement pathway activation. Together, these data point to a previously unrecognised role of the complement system and raise the possibility of drug repurposing of complement-directed therapeutics in GCA.

**Figure 1.**
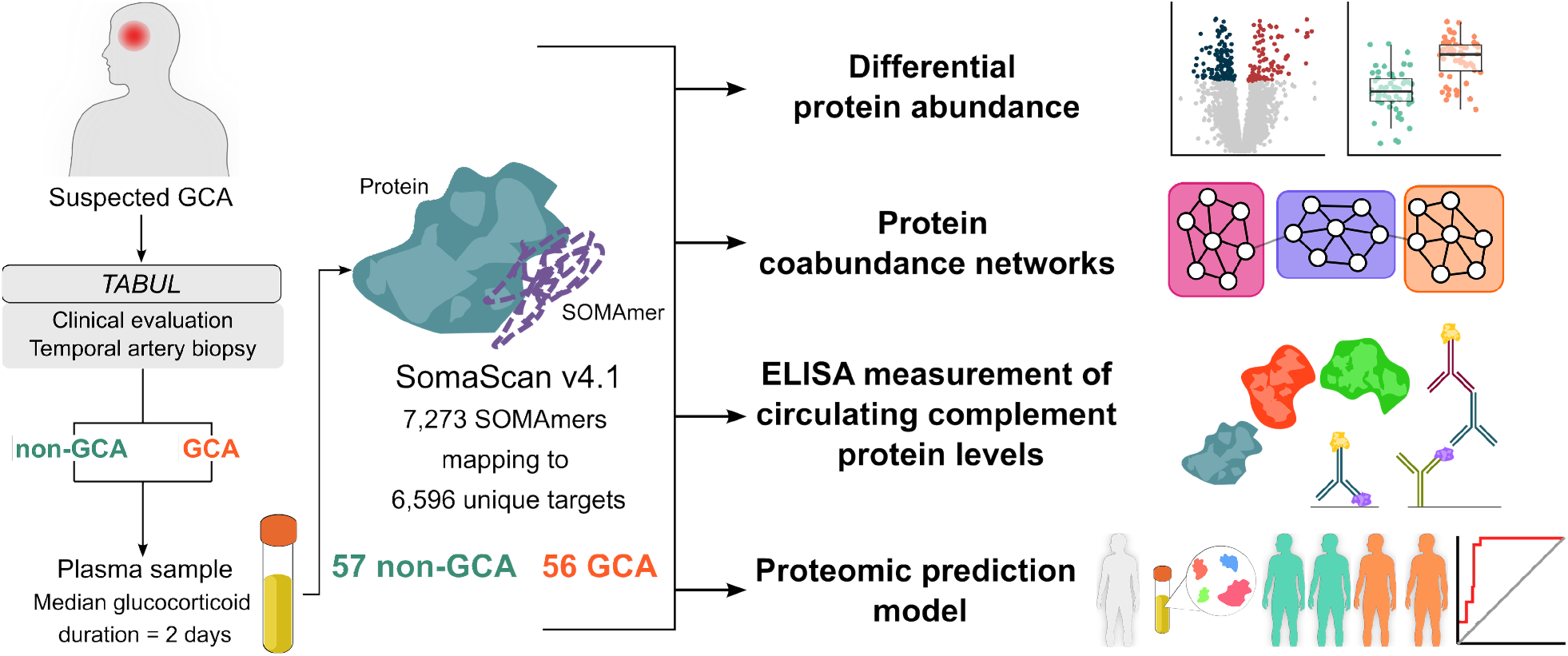
Overview of proteomic data generation and key analysis workflows. Proteomic measurements were performed in a subset of participants from the TABUL study, which recruited patients presenting acutely with possible GCA. In the present study, 57 patients had biopsy-proven GCA and 56 had GCA excluded both clinically and on biopsy (‘non-GCA’). Two samples (1 GCA, 1 non-GCA) were excluded during proteomic quality control. The number of SOMAmers reported reflect those available for analysis following quality-control.

## Results

### Clinical cohort characteristics

Plasma samples were collected at baseline from individuals presenting with possible cranial giant cell arteritis recruited to the TABUL study (10). In TABUL, patients were treated at presentation empirically with glucocorticoids as per standard clinical guidelines and a TAB was performed within 7 days of presentation. Based on the clinical assessment, including longitudinal follow-up data, laboratory tests and TAB, patients were adjudicated to either have GCA or not GCA at 6 months. The TABUL study allowed a clinical diagnosis of GCA despite a negative TAB. However, our previous work showed that this TAB negative GCA group do not share the proteomic signature seen in TAB positive GCA, raising the possibility that TAB negative GCA contains misclassifications (9). To avoid diagnostic ambiguity in the present study, we limited proteomic measurements to samples from patients with biopsy-proven GCA (n = 56) and individuals who did not have GCA (non-GCA; n = 57), defined as both negative TAB and a clinical conclusion that the patient did not have GCA.

Proteomic measurements were performed using the aptamer-based SomaScan v4.1 platform. After quality control (QC), 7,273 measurements (SOMAmers) corresponding to 6,596 unique protein targets were available for 55 GCA samples and 56 non-GCA samples (**Supplementary Figure 1, Supplementary Data 1&2**). Duration of glucocorticoid treatment duration at the time of research blood sampling was short (median 2 days). There were no significant differences in glucocorticoid treatment duration (*P* 0.39, two-sided Wilcoxon rank-sum test), age (*P* 0.64, two-sided Wilcoxon rank-sum test) and reported sex (*P* 0.42, two-sided Fisher’s exact test) between GCA and non-GCA groups.

### Proteomic associations with GCA

To identify proteins associated with GCA, we performed differential protein abundance analysis comparing GCA with non-GCA samples. We refer to proteins by their encoding gene symbol to provide a standardised nomenclature. 295 SOMAmers corresponding to 274 unique protein targets were significantly associated with GCA (*P*_FDR_ < 0.05), with 144 SOMAmers increased and 151 decreased (**Figure 2a, Supplementary Data 3**). Specific proteins differentially abundant in GCA patients reflected pathophysiological processes in GCA including an acute phase response (SAA1, SAA2, CRP, HP, LBP and FGG), cytokines and chemokines (IL36A, PPBP, CXCL2, CXCL3, CXCL9, CXCL13, CCL18, CCL23, CCL25), and distinct aspects of vascular remodelling such as extracellular matrix breakdown (MMP1, MMP12 and TIMP1), neovascularisation (ABGPT1; angiopoietin-1) and fibrosis (CTHRC1 and SERPINH1). Of the GCA-associated proteins, MFGE8 (lactadherin) and PBK (PDZ binding kinase) are encoded by genes located in two of the four non-*HLA* genetic risk loci identified in the largest genome-wide association study (GWAS) to date (11).

**Figure 2.**
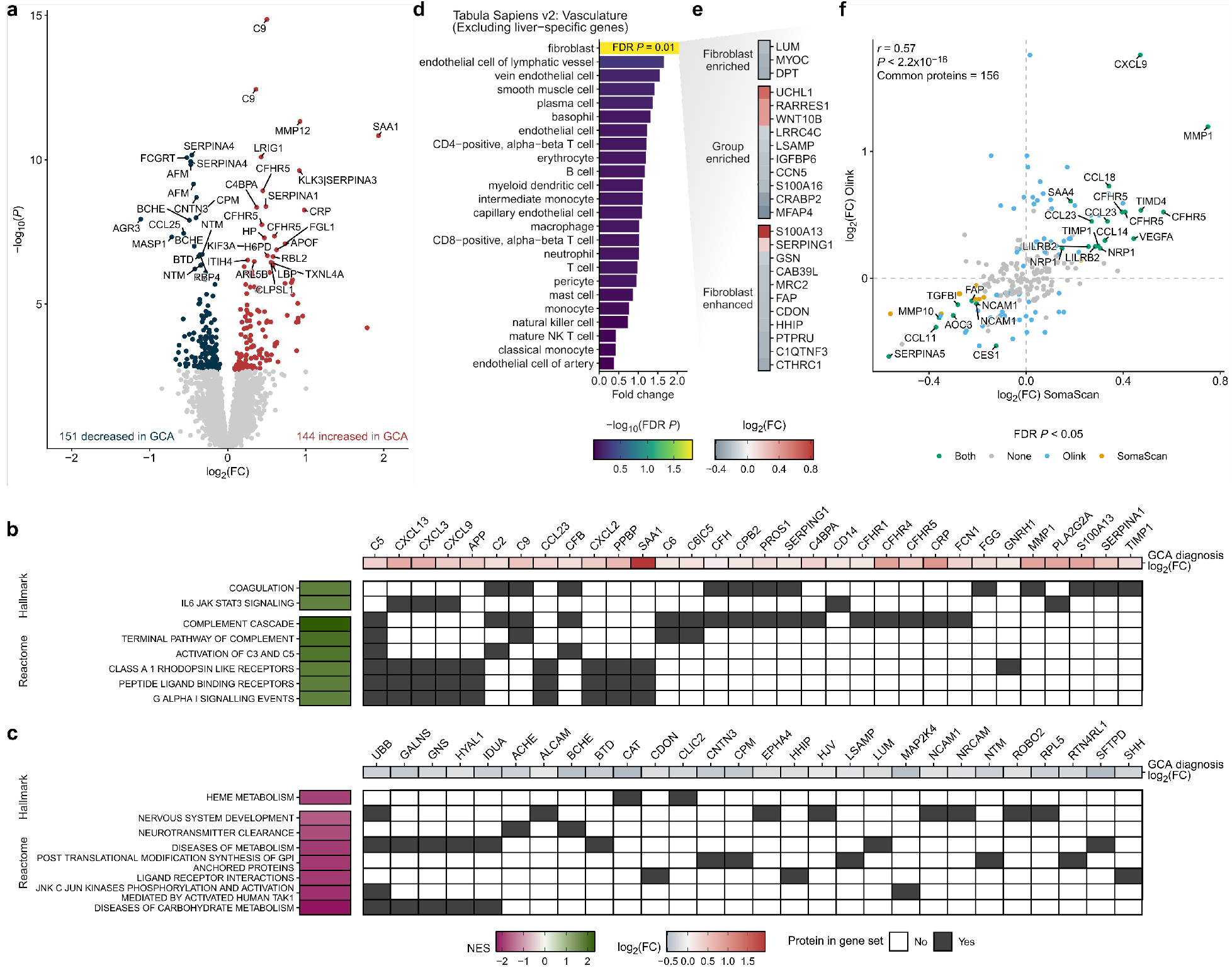
Plasma proteomic changes in giant cell arteritis. Proteins labelled with NCBI gene symbols. **(a)** Differentially abundant proteins in biopsy-proven GCA relative to non-GCA samples. Red (increased in GCA) and blue (decreased) points indicate proteins significantly associated with GCA (*P*_FDR_ < 0.05). FC= fold change. **(b, c)** Gene Set Enrichment Analysis (GSEA) using ranked GCA association moderated *t*-statistics. Reactome and Hallmark gene sets are stratified according to whether they are increased (**b**) or decreased (**c**) in GCA relative to non-GCA. GSEA leading-edge proteins significantly associated with GCA in the differential protein abundance analysis are shown on the upper panels. **d)** GCA-associated proteins are overrepresented in genes specific to vascular fibroblasts. Liver-specific genes were excluded from testing. Vasculature cell-specific genes were identified using Tabula Sapiens v2 single-cell RNA-sequencing data (81). **(e)** GCA-associated proteins encoded by vascular fibroblast-specific genes. Colour gradient key indicates effect size (red; increased in GCA). **(f)** Comparison of effect sizes (biopsy-proven GCA vs non-GCA) for proteins measured using antibody-based (Olink) (9) and aptamer-based (SomaScan) proteomic platforms on the same samples. *P*_FDR_ values calculated based on the number of SOMAmers targeting proteins measured on both platforms to enable fair comparison. *r* = Pearson’s correlation coefficient.

To identify dysregulated biological pathways in GCA, we mapped SOMAmer protein targets to Hallmark and Reactome (12, 13) gene sets and performed Gene Set Enrichment Analysis (GSEA, **Supplementary Data 4**). Upregulated pathways included coagulation, IL6-JAK-STAT3 signalling, complement, G-protein coupled (GPC) receptor signalling (**Figure 2b**). Inspection of leading-edge proteins for the coagulation pathway revealed upregulation of PROS1 (protein S) and FGG (fibrinogen) but also complement proteins, alpha1-antitrypsin (SERPINA1) and matrix metalloproteinases, reflecting non-specificity of the pathway annotation. The GPC receptor signaling pathways were largely characterised by GCA-associated cytokine and chemokines (**Figure 2b**). Conversely, we observed negative enrichment scores for pathways including metabolism (heme metabolism; diseases of carbohydrate metabolism; diseases of metabolism), neurobiology (nervous system development and neurotransmitter clearance) and cellular signalling (Ligand-receptor interactions; post-translational modification: synthesis of GPI-anchored proteins; JNK (c-Jun kinases) phosphorylation and activation mediated by activated human TAK1) (**Figure 2c**). Examination of leading-edge proteins for the ‘diseases of metabolism’ and ‘diseases of carbohydrate metabolism’ pathways pointed to a common theme of lysosomal degradation of glycosaminoglycans and extracellular matrix polysaccharides, perhaps reflecting altered turnover of extracellular matrix glycosaminoglycans.

To infer potential biological sources of plasma proteins dysregulated in GCA, we tested for overrepresentation of GCA-associated proteins in public tissue- and cell-specific gene expression datasets (**Methods**). GCA-associated proteins were overrepresented among liver-specific genes and were more likely to be secreted into blood, likely reflecting acute phase response proteins (**Supplementary Figure 2a, b; Supplementary Data 5, 6**). Within vascular tissue, GCA-associated proteins were overrepresented among genes exhibiting increased specificity to vascular fibroblasts; this held true after excluding proteins largely produced by the liver (**Figure 2d, Supplementary Figure 2c-e, Supplementary Data 7, 8**). Proteins with the highest tier of fibroblast-specificity were reduced in GCA, including the extracellular matrix components LUM (lumican), DPT (dermatopontin) and MYOC (myocilin) (**Figure 2e**). FAP (fibroblast activation protein-α), a marker of activated fibroblasts, was also reduced in GCA. Fibroblast-specific proteins increased in GCA included UCHL1, RARRES1, WNT10B, S100A13 and SERPING1. We did not detect significant overrepresentation of GCA-associated proteins in blood-cell or lymphoid organ specific genes (**Supplementary Figure 2g; Supplementary Data 9**).

In Maughan *et al* (9), we used an antibody-based proteomic platform (Olink Target Inflammation and Cardiometabolic panels) to measure a targeted set of plasma proteins in GCA and non-GCA samples, including all samples in the present study. 156 proteins were measured on the Olink and SomaScan proteomic platforms (represented by 222 SOMAmers since some proteins are targeted by multiple SOMAmers). To evaluate consistency of proteomic associations across platforms, we compared results using SomaScan to those from Olink, re-analysing the datasets to ensure consistent methodology and comparable multiple testing correction (**Methods**). Estimated effect sizes (log_2_ fold changes between biopsy-proven GCA and non-GCA) were broadly consistent across platforms (Pearson’s *r* 0.57, **Figure 2f, Supplementary Data 10**). Unsurprisingly, proteins significantly associated with GCA in both platforms exhibited more consistent effect sizes (Pearson’s *r* 0.85, **Supplementary Figure 3**). 20 proteins were significantly associated with GCA in both platforms, with 12 proteins increased (CFHR5, NRP1, MMP1, TIMP1, TIMD4, CXCL9, CCL14, CCL18, CCL23, LILRB2, SAA4, VEGFA) and 8 proteins reduced (TGFBI, AOC3, SERPINA5, FAP, NCAM1, MMP10, CES1, CCL11), thereby identifying a high-confidence set of proteins robustly associated with GCA (9).

### Plasma protein networks in GCA

Applying Weighted Gene Co-expression Network Analysis (WGCNA) (14) to the proteomic data, we identified 37 modules of co-abundant proteins (24-591 SOMAmers each; **Figure 3a, b; Supplementary Data 11**). Testing module eigenproteins versus clinical traits showed that Modules 19 and 23 were significantly associated with GCA diagnosis and CRP, ESR and platelet counts (*P*_FDR_ < 0.05, **Figure 3c**; **Supplementary Data 12**). Module 8 associated with ESR, while Modules 5, 8 and 15 associated with platelet count, but none with GCA diagnosis (**Figure 3c**). No modules or individual SOMAmers were significantly associated with polymyalgic symptoms or cranial ischaemic complications (**Figure 3c**; **Supplementary Figure 4a, b; Supplementary Data 12**).

**Figure 3.**
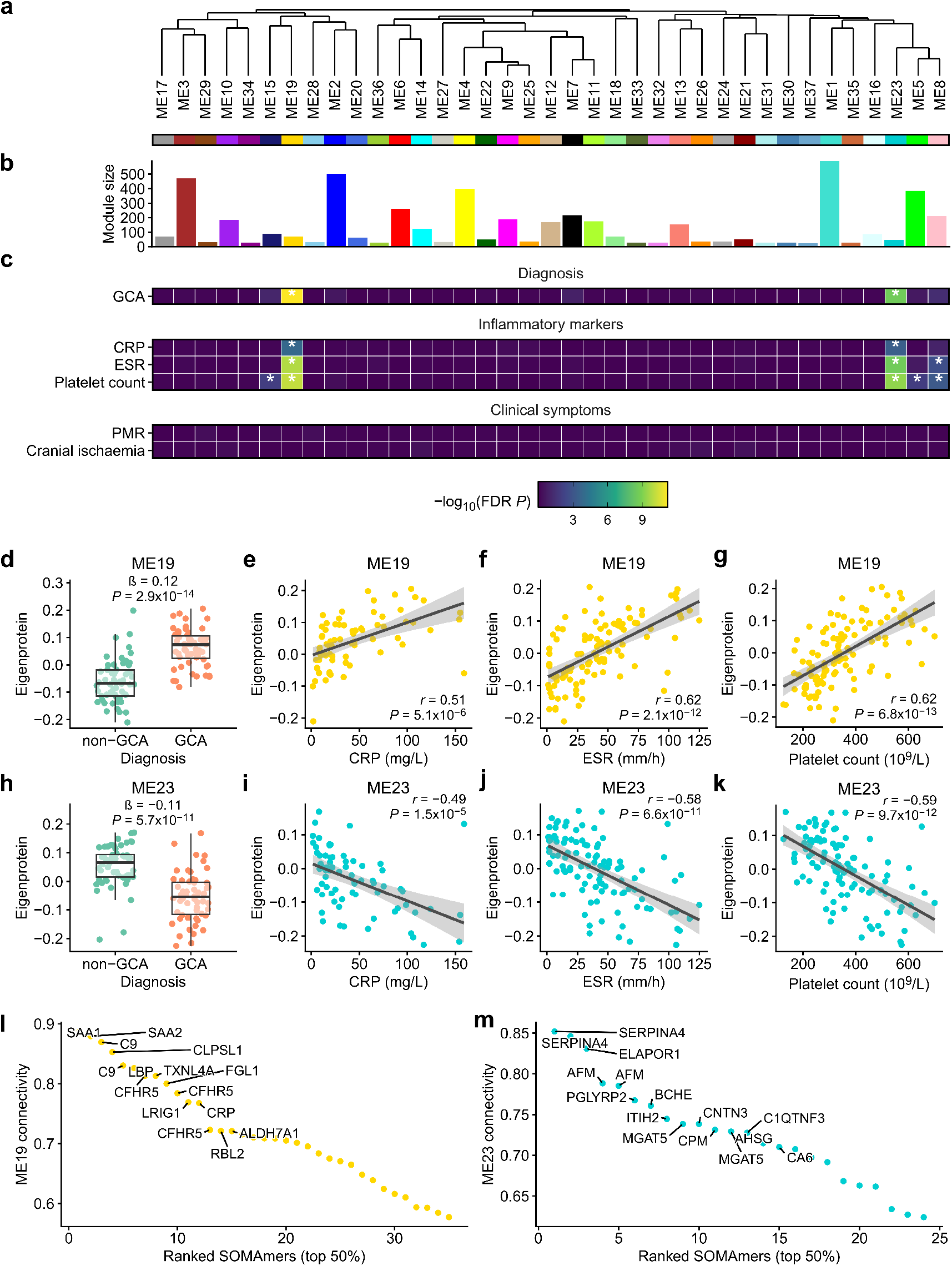
Network analysis identifies protein modules associated with GCA diagnosis and inflammatory markers. **(a)** Hierarchical clustering of protein coabundance modules using a signed dissimilarity matrix of module eigenproteins. **(b)** Number of SOMAmers in each module. **(c)** Association of module eigenprotein values with GCA diagnosis, inflammatory markers and clinical symptoms. *P*FDR values corrected for multiple testing across all module and trait comparisons. * = *P*FDR < 0.05. **(d-k)** Module 19 and 23 eigenprotein associations with: (**d, h**) GCA diagnosis, (**e, i**) CRP, (**f, j**) ESR and (**g, k**) Platelet count. **(l, m)** Connectivity of module 19 (**l**) and 23 (**m**) members. Proteins are labelled by NCBI gene symbol (duplicates reflect multiple SOMAmers targeting the same protein). PMR = Polymyalgia rheumatica, CRP = C-reactive protein, ESR = Erythrocyte sedimentation rate.

Modules 19 and 23 showed opposing associations with clinical traits. Module 19 eigenprotein values were elevated in GCA, and positively correlated with CRP, ESR and platelet count (**Figure 3d-g**), while Module 23 showed the inverse relationship (**Figure 3h-k**). In both modules, proteins with higher network connectivity showed stronger trait associations (**Supplementary Figure 5c-j, Supplementary Data 11, 13-15**). Important Module 19 members included SAA1, SAA2, C9, CLPSL1 and CFHR5 (**Figure 3l**), while important Module 23 members included SERPINA4, ELAPOR1, AFM, PGLYRP2 and BCHE (**Figure 3m**). Overrepresentation analysis linked Module 19 to the complement system, innate immunity and toll-like receptor signalling, whereas no pathways were enriched in Module 23 (**Supplementary Figure 5k, Supplementary Data 16**). Together, these findings highlight coordinated networks of plasma proteins dysregulated in GCA, corresponding to inflammatory read-outs of disease activity, and again implicate complement pathway dysregulation.

### Altered levels of circulating complement proteins in cranial GCA

Given the enrichment of complement pathways in GCA (**Figure 2a, e**), we next examined differences in individual components of the complement system. We manually curated a list of 64 SOMAmers targeting 50 unique components of the complement system (**Figure 4a**, **Supplementary Data 17**). In the lectin pathway, two SOMAmers targeting FCN1 were increased in GCA, while a SOMAmer targeting the MASP3 heavy chain, which is shared with MASP1 (15), was decreased. Factor B, part of the alternative pathway, was increased in GCA. Downstream of complement initiation, intact C2 and C5 were increased along with C5 cleavage fragments, the proinflammatory anaphylatoxin C5a and C5b-C6 complex, primer for the Terminal Complement Complex (TCC). Other potential components of the TCC, C6 and C9 were also increased. Several proteins involved in the regulation of the complement system had altered circulating levels, with increased levels of C1-Esterase inhibitor, the alpha chain of C4b-binding protein and reduced levels of CD46. Factor H, the negative regulator of the alternate complement pathway, was increased, as were several members of the Factor H-related protein family: CFHR1, CFHR4 and CFHR5.

**Figure 4.**
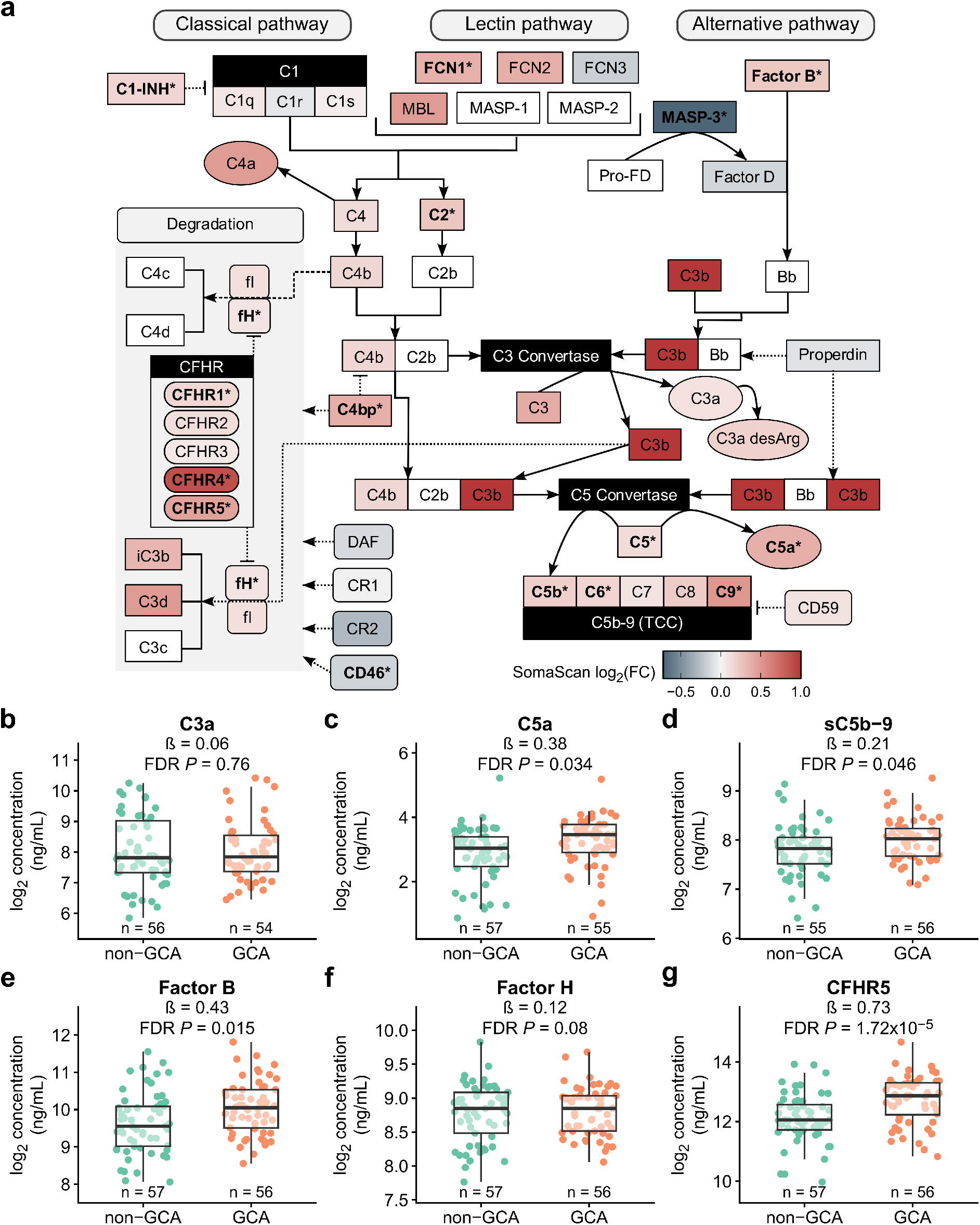
Complement pathway activation in GCA. **(a)** Schematic of the complement system. Proteins are coloured by SOMAmer log2 fold changes comparing GCA to non-GCA samples. Targets in bold followed by an asterisk indicates that protein has an *P*FDR value < 0.05. White boxes indicate proteins not measured by SomaScan v4.1. Where multiple SOMAmers share the same target, the summary statistics for the SOMAmer with the smallest P-value are shown. **(b-g)** ELISA measurements of the complement proteins: C3a (**b**), C5a (**c**), soluble C5b-9 (**d**) and Factor B (**e**), and regulatory proteins Factor H (**f**) and CFHR5 (**g**). The sample size tested for each protein is provided above each study group.

To validate the SomaScan findings, we performed dedicated measurements of complement proteins and their regulators using enzyme-linked immunosorbent assays (ELISAs). In aliquots of the same plasma samples used for SomaScan measurements, we quantified levels of complement fragments C3a, C5a, soluble C5b-9 (sC5b-9, i.e. TCC), Factor B, Factor H and CFHR5. We assessed consistency of SomaScan measured complement proteins against ELISA measurements. Three SOMAmers measuring CFHR5 all showed strong correlation with ELISA measurement (Spearman’s rho 0.65-0.73, **Supplementary Figure 6f-h**). SomaScan-measured C5a (Spearman’s rho 0.59) and Factor B (Spearman’s rho 0.52) also corresponded well to ELISA measurements (**Supplementary Figure 6c, d)**. Factor H (Spearman’s rho 0.32) and C3a (Spearman’s rho 0.21-0.27) measurements were less strongly correlated to their corresponding ELISA values (**Supplementary Figure 6a, b, e**). Correlations of SomaScan-measured components of the TCC against ELISA-measured sC5b-9 ranged from -0.22 to 0.19 (Spearman’s rho, **Supplementary Figure 7**).

We next compared circulating complement activation markers between GCA and non-GCA (**Supplementary Data 18**). C5a and sC5b-9, but not C3a, were significantly increased in GCA (**Figure 4b-d**). We also observed significantly elevated levels of Factor B (**Figure 4e**). We then assessed changes in regulatory complement proteins, observing no difference in Factor H levels (**Figure 4f**), but significantly increased CFHR5 levels in GCA (**Figure 4g**). These findings suggest activation of the terminal complement pathway in GCA.

### A plasma protein signature of GCA

No specific blood-based diagnostic biomarker currently exists for GCA. We therefore sought to develop a multi-protein signature using LASSO logistic regression to distinguish GCA from non-GCA samples (**Figure 5a**). Aggregated model prediction scores on held-out samples across GCA prediction models were significantly higher for GCA than non-GCA samples (median prediction score 0.86 versus -0.93, respectively, **Figure 5b**). Receiver-operator characteristic curve (ROC) analysis showed that GCA samples were distinguished from non-GCA samples (area-under-the-curve (AUC) 0.93, 95% CI 0.88-0.98; **Figure 5c**). We then used all 111 TABUL samples to fit a final GCA prediction model for external validation. Our final GCA prediction model contained 26 predictors, with the most important predictors including MMP12, C9 SYF2, PHF11, FCGRT, KLK3|SERPINA3, CCL25, H6PD, APOL1 and AGR3. (**Figure 5d**, **Supplementary Data 19**).

**Figure 5.**
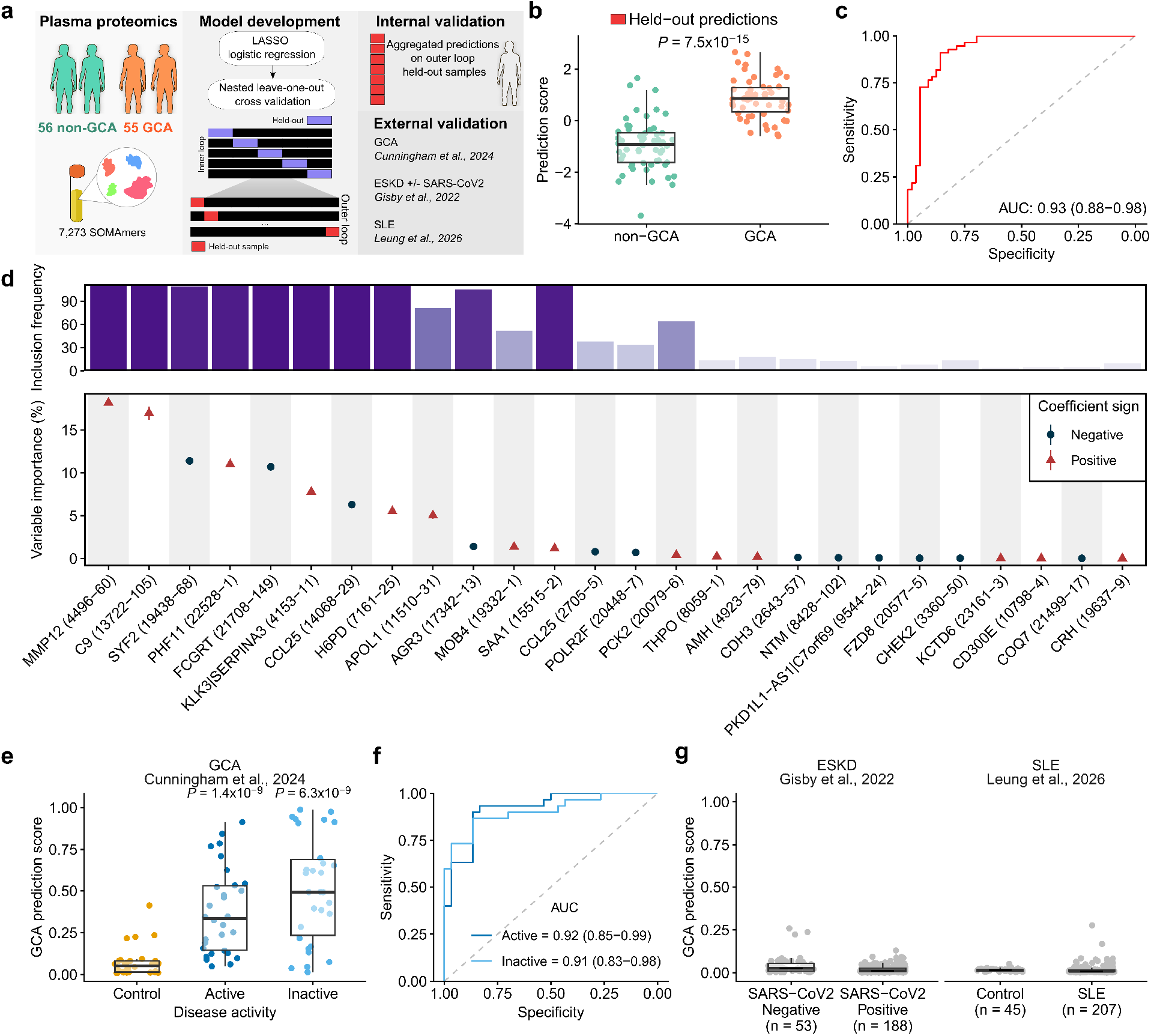
Development of a proteomic GCA prediction model. **(a)** Proteomic GCA prediction model development using a nested cross-validation framework. **(b)** Model predictions on held-out TABUL samples (n = 111) during model development. *P* value from a two-sided Wilcoxon-rank sum test. **(c)** Receiver operator characteristic (ROC) curve for held-out predictions on TABUL samples showing the area under the ROC curve (AUC) with 95% confidence interval. **(d)** Proteomic predictors included in the final GCA prediction model trained on all TABUL samples. Predictors are ordered by mean importance. Error bars show standard error of the mean. The top-hand panel shows the number of models each predictor was included in during model development. **(e)** GCA model predictions on external proteomic data generated by Cunningham *et al* (16). *P* values calculated from a two-sided Wilcoxon ranked-sum test comparing healthy control sample (n = 30) predictions to active (n = 30) and inactive GCA (n = 30) samples and Bonferroni-corrected by two tests. **(f)** ROC curve and AUCs (with 95% CI) for proteomic GCA model predictions distinguishing active and inactive GCA from healthy samples Control samples from Cunningham *et al* (16). **(g)** GCA model predictions for proteomic data from Gisby *et al* (17) (patients with end-stage kidney disease (ESKD) with or without SARS-CoV 2 infection) and Leung *et al* (18) (SLE patients and healthy volunteers).

To externally validate our model, we used publicly available plasma proteomic data (SomaScan v4.0) generated from cranial and large-vessel GCA patients (combined n = 30, initially recruited during active disease and resampled during remission) and controls (n = 30) (16). Model prediction scores were higher in active and inactive GCA samples relative to controls (Bonferroni-adjusted *P* values < 0.05, two-sided Wilcoxon-rank sum test), albeit with an apparent bimodal distribution present (**Figure 5e**). Our GCA prediction model distinguished active GCA (AUC 0.92, 95% CI 0.85-0.99) and inactive GCA samples (AUC 0.91, 95% CI 0.83-0.98) from control samples with comparable ability to our internal validation (**Figure 5f**). Given that our model was trained on cranial GCA patients, we examined whether GCA subtype impacted prediction performance. Our model performed well in distinguishing patients with both cranial GCA (n = 21) and large-vessel GCA (n = 9) relative to controls, across active and inactive disease states (AUCs 0.82-0.96, **Supplementary Figure 8**), suggesting the model captures changes in vascular biology beyond just acute inflammation.

To evaluate the specificity of our GCA prediction model in the context of other inflammatory states, we calculated prediction scores in samples from proteomic datasets (SomaScan v4.1) in which inflammatory plasma protein signatures have been previously reported (17, 18). These datasets included an end-stage kidney disease (ESKD) patient cohort with (n = 53) or without (n = 188) COVID-19 (17); and a cohort comprising SLE patients (n = 207) and controls (n = 40) (18). The median prediction score across the study groups across both cohorts ranged from 0.008-0.026 (**Figure 5g**), indicating the GCA proteomic model is specific and not merely capturing systemic inflammation.

## Discussion

In this study, we performed broad profiling of the plasma proteome in patients presenting with possible cranial GCA. This study design allowed us to compare circulating protein levels in patients with biopsy-proven GCA versus individuals in whom GCA was confidently excluded both clinically and by histopathology. A notable finding was enrichment of the complement pathway in GCA, with complement proteins among the most strongly upregulated proteins. We validated these findings using ELISAs, confirming elevation of plasma C5a, sC5b-9, Factor B and CFHR5. Increased concentrations of Factor B may be the result of increased Factor B synthesis or activation of the alternative complement pathway. Elevated C5a and sC5b-9 provide strong evidence of complement terminal pathway activation.

A role for complement has not been previously appreciated in GCA. The complement pathway has long been recognised to play a role in immune complex-mediated diseases such as SLE. More recently, its importance has been recognised in AAV, with mouse models demonstrating the importance of C5a in MPO-ANCA mediated crescentic glomerulonephritis (19, 20). C5a is a potent anaphylatoxin and chemoattractant for cells expressing its cognate receptor C5aR1. This work led to the development of avacopan, a C5aR1 receptor inhibitor. In AAV, the mechanism of action of avacopan is thought to primarily be inhibition of neutrophil chemotaxis and activation. Although avacopan has recently been withdrawn in some jurisdictions due to concerns regarding the conduct of the ADVOCATE trial (publication retracted) (21), post-marketing experience suggests this may be an effective drug (22). In GCA, unlike AAV, neutrophilic infiltrates are not a prominent histopathological feature, although a role for neutrophils in pathogenesis has been proposed (23). NETs have been identified in the adventitia around the vasa vasorum (24) and *CCDC25*, the gene encoding the NET receptor, lies in a GCA susceptibility locus (11). Alternatively, C5a may play in a role in GCA through effects on cell types other than neutrophils. C5aR1 is also expressed on monocytes, macrophages and dendritic cells (DCs) and C5a can attract and activate monocytes and macrophages. In addition, activation of C5aR1 on dendritic cells (DCs) affects proliferation and differentiation of naïve T-cells (25). Our findings raise the possibility that avacopan or other C5a-targeted therapies could be re-purposed for GCA.

In GCA there was strong enrichment of the IL6-JAK-STAT3 pathway. Upadacitinib, a JAK inhibitor, has recently demonstrated efficacy in GCA (8), illustrating the potential of proteomics to highlight therapeutically important pathways. Consistent with previous studies and the known importance of IL6 in GCA, we identified increased levels of circulating acute phase reactants such as C-reactive protein (CRP) (26, 27, 3) and serum amyloid A (28–30), alongside increases in Haptoglobin (HP), Lipopolysaccharide binding protein (LBP) and the gamma chain of Fibrinogen (FGG). Notably, SOMAmer-measured IL-6 was not significantly elevated with GCA in this study, despite a large body of literature supporting this association (31–34), including our previous data generated on Olink proteomic platform in the same set of samples (9). We and others (35) have previously noted attenuated associations when analysing SomaScan-measured IL-6 compared to Olink measured IL-6 in other datasets, including in COVID-19 (36, 17), where elevation of IL-6 levels is very well established (37–39), suggesting that the SOMAmers measuring IL-6 perform poorly.

We observed a complex chemokine signature indicating activation of both innate and adaptive immunity. CXCL2 and CXCL3 act on CXCR2⁺ myeloid cells, particularly neutrophils, and CCL23 acts on CCR1_+_ cells, particularly monocytes. Upregulation of CCL18 is consistent with a macrophage/myeloid-driven inflammatory programme, as previously described in AAV (40). CCL25 is expressed predominantly by thymic and intestinal epithelial cells and directs the trafficking of CCR9⁺ lymphocytes and subsets of dendritic cells. Its cellular source and functional significance in GCA are thus unclear. CXCL9 and CXCL13 act on adaptive immune cells. CXCL13 recruits CXCR5⁺ cells (particularly B cells but also Tfh cells) and is associated with ectopic/tertiary lymphoid organ (TLO) formation in chronic inflammation. Arterial TLOs have been described in GCA with increased *CXCL13* expression (41). Together these data highlight a potential role for B cells in GCA pathogenesis, although since these data are observational, they do not prove causality.

Strongly elevated plasma CXCL9 in GCA was a consistent finding in our data across both SomaScan and Olink platforms and concordant with previous reports (42–44, 9). CXCL9 is potently induced by interferon-γ (IFN-γ) (45–47), which is produced by Th1 cells that are both expanded in the circulation and temporal arteries in GCA (48). IFN-γ acts on vascular smooth muscle cells (VSMCs) within the artery wall to induce release of CXCL9 and related chemokines, which in turn recruit CXCR3-expressing monocytes, T-cells and B cells into the vessel wall, establishing a feed-forward loop of chemokine-driven immune cell recruitment. A mechanistic link is supported by a study of cultured temporal arteries from GCA patients, in which IFN-γ blockade directly reduced CXCL9, CXCL10, and CXCL11 expression and decreased macrophage infiltration (49). Elevated circulating CXCL9 has also been detected prior to clinical GCA diagnosis (50), suggesting that this chemokine response is an early event in disease pathogenesis rather than merely a downstream consequence of established arteritis. Together, these findings support circulating CXCL9 as a candidate biomarker of GCA-specific vascular pathology that may offer discriminative value independent of general systemic inflammatory markers. The prominent elevation of CXCL9 raises the possibility as to whether therapeutic inhibition of IFNγ might be a potential therapeutic strategy in GCA. Emapalumab, an anti-IFNγ monoclonal antibody, has been developed for haemophagocytic lymphohistiocytosis (HLH); however, inhibition of a key facet of the anti-viral response in an elderly GCA population might lead to unacceptably high infectious complications.

Although we studied circulating proteins in blood, we observed evidence of arterial tissue-derived proteins indicative of vascular injury and remodelling. The direct contact between the vasculature and the circulation likely make plasma proteomics a valuable read-out of arterial tissue pathology. Proteins associated with a GCA diagnosis were overrepresented in genes specific to vascular fibroblasts. The role of fibroblasts and their interactions with immune cells in GCA pathogenesis is increasingly appreciated, with fibroblasts contributing to the extracellular matrix maintenance, proinflammatory cytokines release, and vascular remodelling via differentiation into myofibroblasts (51). GCA-associated proteins with vascular fibroblast specificity were generally reduced in GCA patients, which may reflect tissue sequestration. As an example, we identified reduced plasma levels of FAP in GCA, consistent with previous reports (9, 52), yet increased expression of *FAP* in temporal arteries of GCA patients has been reported (52).

We observed increased plasma levels of the matrix metalloproteases MMP1 and MMP12 and TIMP1 (a matrix metalloproteinase inhibitor) and reduced levels of MMP10. These findings were consistent across both the SomaScan and Olink platforms, indicating that they are robust. Matrix metalloproteinases (MMPs) are produced by macrophages, vascular smooth muscle cells and myofibroblasts, contributing to tissue injury in GCA, including degradation of the internal elastic lamina. Increased circulating levels of MMP12 and MMP1 have been reported by other groups (53, 54). In temporal arteries from GCA patients, studies have shown increased MMP12 transcript levels and the colocalization of MMP12 in macrophages and giant cells (54–56). Previous studies have reported elevated MMP2 and MMP9 in GCA (57, 58), but there were no significant differences in these MMPs in our data. A previous study reported increased circulating TIMP1 levels in active relative to inactive GCA (59). These findings demonstrate the ability of plasma proteomics to reflect not only systemic immune responses but also tissue pathobiology.

Neoangiogenesis in GCA provides a conduit for infiltrating immune cells and is an important factor in disrupting the normally immune-privileged artery wall. We observed elevated plasma VEGF-A in both the SomaScan and Olink data, in line with previous studies (60). New vessel formation has been shown to correlate with increased local vascular endothelial growth factor (VEGF-A) expression (61). We also observed increased plasma ANGPT1 (angiopoietin-1). In a previous study, serum angiopoietin-1 levels were not altered in acute untreated GCA patients compared to healthy controls (62), yet within GCA, higher levels predicted a shorter time to glucocorticoid-free remission, potentially consistent with protective effects on vasculature function (63). We found lactadherin, a secreted glycoprotein implicated in VEGF-mediated neovascularisation (64), reduced in GCA. A GCA GWAS with fine mapping identified an intronic variant (rs11073821, GRCh38:15:88903861) within the gene encoding lactadherin (*MFGE8*), as a candidate causal variant (11). The risk allele (A) is associated with reduced SomaScan-measured lactadherin abundance in plasma (65, 66) and reduced *MFGE8* expression in T-cells (67) and fibroblasts (68). In GCA, *MFGE8* expression was also reduced in TAB with transmural inflammation compared to biopsies with adventitia-limited inflammation (69).

We developed a multi-protein model that could accurately identify GCA. At present, the diagnosis of GCA relies on imaging (usually temporal artery and axillary ultrasound) and/or TAB. Sensitivity of both modalities falls after initiation of glucocorticoid therapy, with US sensitivity particularly time dependent. In some healthcare settings, there may be logistical difficulties in arranging rapid US or TAB. For TAB, there is a risk of false negatives from ‘skip lesions’ which are normal segments of arteries that lie between inflamed regions. Thus, the situation can arise where the US and TAB are negative, but the clinician does not feel confident that GCA can be reliably excluded, resulting in patients committed to prolonged steroid treatment. A protein-based biomarker taken at the time of initial presentation could help address such diagnostic ambiguity and would also be valuable in acute care settings to triage patients who require further investigations from those in whom GCA can be confidently excluded.

Our model was developed in individuals presenting with symptoms that could indicate cranial GCA, which reflects the real-life decision making encountered in acute healthcare settings where such a model would be most useful. To develop the model, we employed a nested leave-one-out cross validation framework to minimise bias when training parameters and to make full use of the sample size available (70). Internal validation showed our model was able to distinguish GCA participants from non-GCA participants with comparable performance metrics to those reported for imaging techniques (71). Our final model, containing 26 protein predictors, successfully generalised to an external SomaScan proteomic dataset (16) generated on GCA patients and healthy controls, with similar performance to our internal validation. The model also appeared to generalise to patients with large-vessel GCA in the external dataset, although the small sample size for large-vessel GCA (n = 9) means that further evaluation is required. Notably the model performed well in predicting both active and inactive GCA, despite having been developed using samples obtained during acute presentations. This suggests that the model may capture aspects of underlying disease biology that persist beyond periods of overt inflammation and may therefore be less sensitive to timing than current tests such as ultrasound. Moreover, it correctly predicted that patients with other diseases characterised by prominent systemic inflammation (SLE and COVID-19) did not have GCA. Together, these findings suggest that the model captures a proteomic signature specific to GCA pathology rather than non-specific inflammation. Consistent with this interpretation, circulating levels of MMP12, the most important predictor in our model, have previously been shown to be unaffected by IL-6 pathway inhibition (54, 72).

Further work is needed to refine the biomarker for clinical translation. While the proteomic GCA prediction model we present here shows the potential of circulating multi-protein biomarkers in the investigation of GCA, further studies are needed to evaluate the signature in multiple independent cohorts, ideally including other mimics and types of vasculitis. For optimal clinical utility, a parsimonious signature is desirable, and absolute protein quantification is needed. Other desirable features are the inclusion of protein biomarkers that are robust to sample handling and relatively insensitive to the first week of glucocorticoid treatment. Ultimately, a multiplex proteomic biomarker, perhaps combined with clinical scoring systems, might be sufficient to rule out GCA in patients with low clinical probability of GCA, and thus help avoid unnecessary investigation and glucocorticoids in low-risk individuals.

This study contains some limitations. Firstly, our data is observational so cannot separate protein drivers of GCA from downstream responses. Second, all participants were receiving glucocorticoids at sampling, which can alter circulating protein levels. The recruitment of treatment naïve individuals with suspected GCA is difficult as glucocorticoid treatment is recommended as soon as there is a clinical suspicion of GCA (2). Despite this, blood sampling was performed rapidly after commencing treatment (median glucocorticoid treatment duration two days), enabling assessment of disease biology close to the untreated state. Third, we performed a cross-sectional analysis of the plasma proteome which provides a snapshot of the proteome in acute, active disease. Future studies employing longitudinal sampling will be useful for understanding the temporal disease process, particularly given the high rate of relapse in GCA. Finally, as all individuals were recruited with suspected GCA, our study is not directly equivalent to studies which compared GCA to healthy controls. The non-GCA group may contain participants with infectious, inflammatory or vascular diseases which may reduce proteomic differences compared to analysis of GCA versus healthy controls.

In summary, we performed wide-angled plasma proteomic profiling in patients presenting acutely with possible GCA. In GCA, we identified terminal complement pathway activation and validated this with orthogonal methods. Our findings raise the possibility of therapeutic targeting of the complement system in GCA. We present encouraging evidence that a multi-protein blood-based biomarker may help in the diagnostic work-up of GCA.

## Methods

### Sex as a biological variable

Plasma samples were collected from both male and female participants. 73.7% participants were female (**Supplementary Data 1**), reflecting the female preponderance of giant cell arteritis. Sex was included as a covariate in statistical models.

### TABUL cohort

Individuals with suspected cranial GCA were enrolled in The Role of Ultrasound Compared to Biopsy of Temporal Arteries in the Diagnosis and Treatment of Giant Cell Arteritis (TABUL) study (ClinicalTrial ID: NCT00974883) (10). Reference GCA diagnoses (GCA or non-GCA) was defined at six months based on presenting clinical features, TAB results, the 1990 American College of Rheumatology classification criteria, clinical course during the follow-up period, final consultant diagnoses, and verification by an expert review panel, as described previously (73). Citrated blood samples were centrifuged at 2,000g for 15 minutes within 1.5 hours of collection, with the resulting plasma stored at -80°C. For proteomic measurements, we selected all available plasma samples for proteomics for patients with both a reference diagnosis of GCA and a positive temporal artery biopsy (n=56) and for patients in whom GCA was excluded (non-GCA; n=57). Demographic and clinical information of participants is provided in **Supplementary Data 1**.

### Plasma proteomics

Circulating plasma protein levels were measured using the SomaScan v4.1 platform (SomaLogic, Boulder, USA). The SomaScan platform uses microarray hybridisation to measure the levels of 7,596 Slow Off-Rate Modified Aptamers (SOMAmers) following binding to specific protein targets (74). SomaLogic performed routine data standardisation to normalise hybridisation readouts, normalise readouts among different protein dilution groups, reduce variation between plates and individual SOMAmer reagents. Samples were processed across two plates. Proteins levels are measured as relative fluorescence units (RFU) normalised against an internal SomaLogic reference (Adaptive Normalization by Maximum Likelihood; ANML), which we then log_2_ transformed.

### Proteomic data preprocessing and quality control

One sample (non-GCA) was excluded prior to analysis due technical issues during SomaScan processing. 7,273 SOMAmers (6,408 unique UniProt IDs and 6408 unique NCBI gene IDs) remained after quality control and removing non-human SOMAmers. To identify sample outliers, we calculated protein abundance principal components using *prcomp* (R stats v4.6.1), flagging samples ±5 standard deviations (SDs) from the mean PC1 or PC2 scores. Samples ±5 SDs from the per-sample mean interquartile range (IQR) or median protein levels were also flagged. These methods identified one sample outlier (GCA, **Supplementary Figure 1a, b**). The estimated limit of detection (eLOD) for each SOMAmer was calculated using the *calc_elod()* function from SomaDataIO v6.6.1 (https://github.com/SomaLogic/SomaDataIO) using blank buffer samples on each plate as a measure of the background signal. >99% of SOMAmers measured in each sample had protein levels above the eLOD (**Supplementary Figure 1c**).

### Explainable variation in protein abundance

After removal of samples failing QC, protein abundance principal components were recalculated to assess the global structure of proteomic variation. GCA diagnosis was significantly associated with PC2 (**Supplementary Figure 9a, b**). To identify further contributors to global protein abundance variation, we assessed the association between the first 10 proteomic PCs (41.42% cumulative explained variance) and measurable sources of variation (recorded sex, age, SomaScan processing plate, glucocorticoid treatment duration, sample storage time and summary statistics reflecting per-sample global signal intensity) using univariate linear regression. PC1 was associated with per-sample global signal intensity summary statistics (**Supplementary Figure 9c**), with the strongest association between PC1 and the sample interquartile range (IQR) of log_2_ transformed protein levels (R_2_ = 0.74). The contributions of GCA diagnosis, age, sex and the sample IQR of log_2_ protein levels to individual protein abundance variance were also measured using variancePartition v1.42.0 (75), noting the contributions of sources other than GCA diagnosis to protein abundance variation (**Supplementary Figure 9d**).

### Proteomic associations

Associations between protein levels and GCA diagnosis (GCA vs non-GCA) were assessed with linear models implemented in limma v3.68.4 (76). Associations were adjusted for age, sex, glucocorticoid treatment duration (days) and the per-sample interquartile range of log_2_ transformed protein levels (to adjust for systematic differences in overall signal intensity across samples). The second-degree orthogonal polynomials of age, glucocorticoid treatment duration and protein measurement dispersion were included in regression models to account for both linear and non-linear relationships with protein abundance (77). Protein level correlations with CRP, ESR and platelet count were measured with Spearman’s rank correlation. Within-cases analyses included testing for proteomic associations with the presence of symptoms consistent with polymyalgia rheumatica (PMR, yes or no) and reported cranial ischaemic complications (yes or no). Moderated *t*-statistics were calculated using the limma trend method. *P* values were adjusted for multiple testing using the Benjamini-Hochberg procedure and reported as *P*_FDR_ values.

### Gene set enrichment analysis

Gene set enrichment analysis (GSEA) was performed using fgsea v1.38.0 (78). Gene sets were downloaded from Reactome (13) and Hallmark (12) using msigdbr v26.1.0. When multiple SOMAmers mapped to the same NCBI gene ID, the SOMAmer with the highest absolute moderated *t*-statistic was retained. When individual SOMAmers were mapped to multiple gene IDs, we retained SOMAmers if all the gene IDs could be assigned to the same gene set. SOMAmers were ranked in descending order based on moderated *t*-statistics as input for GSEA. Enrichment *P* values were calculated using 10,000 permutations and adjusted for multiple testing across within each gene set resource using the Benjamini-Hochberg procedure.

### Cell-type, tissue and secretome enrichment

The cell-, tissue- and secretome-enrichment of genes encoding GCA associated proteins (*P*_FDR_ < 0.05) was assessed using a background of all genes encoding proteins targeted by SOMAmers. Duplicate SOMAmer targets (i.e. mapping to the same Ensembl gene ID) were pruned by retaining only the SOMAmer with the highest absolute GCA association *t*-statistic. Cell-type and tissue enrichment was assessed using a hypergeometric test implemented in TissueEnrich v1.32.0 (79). Enrichment *P* values were adjusted for multiple testing using the Benjamini-Hochberg procedure and reported as *P*_FDR_ values.

Cell type and tissue specificity was defined according to the Human Protein Atlas (HPA) criteria (80). Tissue-enrichment was assessed using a reference consensus RNA expression resource implemented in TissueEnrich v1.32.0 (79), comprising bulk RNA-sequencing data from HPA and Genotype-Tissue Expression (GTEx) consortia (80, 68). Cell type specificity was assessed using reference single-cell RNA-sequencing (scRNA-seq) data generated on whole blood (85,233 cells) and vasculature (42,650 cells) samples as part of Tabula Sapiens v2 (81). Publicly accessible processed and post-QC scRNA-seq h5ad files were downloaded from CELLxGENE (see **Data Availability**) and converted into SingleCellExperiment objects using anndataR v1.2.0 (82) and SingleCellExperiment v1.34.0 (83). Gene cell type specificity was assigned following an approach modelled on the processing steps of the HPA Single Cell Type Atlas(84). Total decontX (85) corrected counts per protein-coding gene in each cell type were summarised using summarizeAssayByGroup implemented in scuttle v1.22.0 (86). Gene counts were normalised by the total gene counts per cell-type multiplied by 1 million to give the protein coding gene expression per million (pTPM). Per sample trimmed mean of M (TMM) normalisation factors were calculated using edgeR v4.10.1(87) to enable comparisons across cell types. pTPM values were divided by the sample TMM normalisation factors resulting in normalised TPM (nTPM) gene expression estimates. Cell type specificity was then assigned using the HPA criteria on nTPM values (84).

Protein secretome annotations were collected from HPA v24 (downloaded 13_th_ May 2025) (88). Overrepresentation of GCA associated proteins in secretome categories was assessed using a hypergeometric test implemented in the *fora* function from fgsea v1.38.0. *P* values were adjusted for multiple testing using the Benjamini-Hochberg procedure and reported as *P*_FDR_ values.

### Cross proteomic platform concordance

184 plasma proteins were previously assayed in TABUL samples using the proximity extension assay (antibody) based Olink Target *Inflammation 1* and *Cardiometabolic* panels (9). Summary statistics were available for 167 proteins assessing differences in Olink-measured protein levels in biopsy-confirmed GCA (n = 56) compared to non-GCA samples (n = 89). All 111 samples profiled by SomaScan were included in this dataset. Proteins targeted by both Olink and SomaScan panels were identified by matching Ensembl gene IDs. 156 Ensembl gene IDs were present on both panels, corresponding to 222 SOMAmers (156 unique UniProt IDs). To ensure consistency with Maughan *et al*, we repeated differential abundance analysis by fitting a linear model to each SOMAmer regressing diagnosis group against log_2_-transformed protein levels with no covariate adjustment. Linear regression *P* values for 222 SOMAmers were then adjusted for multiple testing using the Benjamini-Hochberg procedure. Log_2_ fold change differences were compared across platforms using Pearson’s correlation.

### Protein coabundance modules

Protein coabundance modules were identified using Weighted Gene Coexpression Analysis (WCGNA) v1.74 (14). Log_2_-transformed protein levels were regressed against the same covariates used in differential abundance analysis. A soft-thresholding power of six was the lowest power with an appropriate scale-free topology fit (R_2_ > 0.85, **Supplementary Figure 5a, b**). A signed network was constructed using the blockwiseModules function. Briefly, a protein coabundance network was constructed using Pearson’s correlation of protein abundance. Proteins underwent average linkage hierarchical clustering, with coabundance modules containing at least 20 proteins identified in the resulting dendrogram with a deepSplit value of 3. Modules were refined by removing proteins with a module connectivity <0.3. Modules were additionally refined by calculating a module dissimilarity matrix and then using a dendrogram cut height of 0.15 to merge modules on that branch.

Biological definitions for protein coabundance modules were assigned by testing overrepresentation of module members in Reactome gene sets using a hypergeometric test implemented in the *fora* function from fgsea v1.38 (78). For this, SOMAmer targets within each coabundance module were mapped to NCBI gene IDs and compared against a background of all NCBI gene IDs targeted by SOMAmers.

Protein coabundance module were tested for association using univariate linear regression of module eigenprotein values (the first principal component of protein members’ abundance) against clinical traits. Traits tested included GCA diagnosis (GCA vs non-GCA), CRP (mg/L), ESR (mm/h) and platelet count (10_9_/L). Other traits tested were restricted to GCA patients only including clinical symptoms, such as whether the individual exhibited symptoms consistent with PMR, or developed cranial ischaemic complications. Linear regression *P* values were adjusted for multiple testing across all modules and traits tested using the Benjamini-Hochberg method.

### ELISA measurements of complement targets

Specific complement targets were measured by ELISA in matched plasma aliquots from 113 TABUL participants (56 GCA, 57 non-GCA). C3a, C5a and soluble C5b-9 complex (sC5b-9) were quantified using MicroVue™ EIA ELISA kits (QuidelOrtho, USA) following the manufacturer’s instructions. Collected plasma was diluted in specimen diluent C5a (1:20), sC5b-9 (1:10) and C3a (1:200). All samples and standards were tested in duplicate.

In-house sandwich ELISAs were used to quantify Factor B, Factor H and CFHR5. Unless stated otherwise, 96-well Maxisorp plates were coated with capture antibody overnight at 4°C in NaHCO₃ buffer (pH 9.5, 0.1M) (Sigma-Aldrich, US). Plates were washed once with phosphate buffered saline (PBS) + 0.1% Tween 20 (Sigma-Aldrich, USA) (PBST) which was used for all subsequent washes. Plates were then blocked with blocking buffer 1% (5% for factor B) bovine serum albumin (BSA; Sigma-Aldrich, USA) in PBST for 1 hour at room temperature. They were then incubated with diluted plasma samples and standards applied in duplicate for 1 hour at room temperature. Plates were washed three times before the detection antibody was applied and another three times before application of secondary antibody. All antibodies and samples were diluted in blocking buffer. Plates were developed using 3,3′,5,5′-Tetramethylbenzidine (TMB) substrate (BD Biosciences, USA). All plates were read at an absorbance of 450nm using a Thermofisher multiscan FC plate reader and SkanIt software v7.0.2 (Thermofisher Scientific, USA). Antibody and standard curve information for in-house sandwich ELISAs can be found in **Supplementary Data 20**.

A small number of samples that had levels outside (above) the range of the standard curve of the ELISA kits. These were repeated at higher dilutions but were clear outliers on QQplots and by Z-scores. These samples were excluded due to the possibility of ex vivo complement activation following sample collection; three samples were excluded for C3a (GCA = 2, non-GCA = 1), two for sC5b-9 (2 non-GCA) and one for C5a (GCA). Protein concentrations were log_2_ transformed prior to downstream analysis. Concordance between SOMAmer and ELISA measured protein abundance for equivalent targets was measured with Spearman’s rank correlation. Differences in ELISA target concentrations between GCA and non-GCA study groups were assessed using linear regression, adjusting for ELISA plate, age, sex and glucocorticoid treatment length. Linear regression *P* values for the six ELISA targets were adjusted using the Benjamini-Hochberg procedure and reported as *P*_FDR_ values.

### Proteomic GCA prediction model development and validation

Log_2_ transformed protein levels were used as predictors of GCA diagnosis (GCA versus non-GCA). Regularised logistic regression models were created using glmnet v5.0 (89), utilising a nested cross-validation framework with nestedcv v0.8.0 (70). Leave-one out cross-validation was implemented on the nested outer loop, whereby each sample acts as the test dataset until all samples have been used, maximising the size of the training dataset whilst allowing assessment of model performance on unseen data. The inner fold was then split into five cross-validation folds for selecting the largest *λ* value within one standard error of the minimum cross-validated error. The α value was set to one to use LASSO regularisation. A prediction model was trained on all training data in the outer loop with predictions made on the held-out sample in the outer loop. This process was repeated until each sample had acted as the held-out sample, creating 111 prediction models. The predictions from all 111 models on each held-out sample were aggregated. Model performance was measured by the C-statistic derived from calculating the area under the ROC curve using pROC v1.19.0.1 (90) on the aggregated predictions. 95% confidence intervals of the C-statistic estimate were calculated using Delong’s method. A final round of 5-fold cross validation on the entire TABUL proteomics dataset produced a final model for external validation.

The final GCA proteomics prediction model was externally validated using publicly available data generated using the SomaScan platform. Cunningham *et al* generated plasma proteomic data on 30 GCA patients (longitudinally sampled during active disease and when in clinical remission) and 30 controls using the SomaScan v4.0 platform (16). Gisby *et al* used the SomaScan v.4.1 platform to generate proteomic data plasma samples from end-stage kidney disease patients with a positive or negative SARS-CoV2 infection (17). Leung *et al* generated plasma proteomics on SLE patients and controls using the SomaScan v4.1 platform (18). We used protein measurements taken from Batch B comprising 207 SLE patients and 45 controls.

### Statistical analysis

All analyses were performed in R v4.6.1. Statistical tests are described in the relevant text and figure legends. Boxplots show the median, with lower and upper hinges showing the first and third quantiles respectively, and whiskers extending to the smallest and largest values no further than 1.5 times the interquartile range from the hinge. Where applicable, multiple testing correction was applied using the Benjamini-Hochberg procedure, with statistical significance defined as an *P*_FDR_ < 0.05.

### Study approval

Ethical approval for the TABUL study [ClinicalTrials.gov: NCT00974883] was granted by the Berkshire Research Ethics Committee (09/H0505/132). Samples are currently held in the Vasculitis and Inflammation Integrated Research Tissue Bank at The University of Leeds which was approved by Wales Research Ethics Committee 4 (20/WA/0195).

## Data Availability

All summary-level data produced in the present study are contained in the manuscript. Individual-level proteomics data have been deposited in Zenodo and will be made publicly available upon journal publication of the final peer reviewed version of the manuscript.

https://zenodo.org/records/23339306

## Author contributions

Study conceptualisation: AWM and JEP. Data curation: LS, RTM, AC, RAL and AWM. Project administration: LS, RTM, AWM, JEP. Methodology: BJW, HLB, RTM, MCP, AWM and JEP. Investigation: BJW, MW, HLB, RTM, MP, MCP, AWM and JEP. Data analysis: BJW, MW, HLB. Visualisation: BJW and MW. Resources: LS, AC, RAL, MCP, AWM, JEP. Supervision: MCP, AWM and JEP. Funding acquisition: MCP, AWM and JEP. Writing (original draft): BJW, MW and JEP. Writing (review and editing): MCP and AWM. All authors read and approved the manuscript.

## AI disclosure

The authors declare that no AI tools were used in the original research for or writing of this article.

## Conflict of interest

A.W.M previously received a research grant from Roche PLC, for unrelated work in GCA, and has undertaken consultancy or received honoraria for speaking at educational events on behalf of her institution from AstraZeneca and Vifor in the last 5 years. R.A.L has received a research grant from Pfizer Global for unrelated work in rheumatology and from UCB, for unrelated work in GCA, and has received honoraria for undertaking consultancy for Amgen, GSK and CSL Vifor; received royalties income for licensed use of disease activity scores in a range of pharmaceutical trials unrelated to GCA; received institutional income for providing training in vasculitis assessment for GSK and CSL Vifor in the last 5 years.

## Funding

This study was supported in part by a Medical Research Foundation award to J.E.P (MRF-042-0001-RG-PETE-C0839), the NIHR Imperial Biomedical Research Centre (BRC) and the NIHR Leeds BRC (NIHR203331). The TABUL study was funded via an NIHR Heath Technology Assessment grant. A.W.M, A.C and L.S were supported by the NIHR Leeds BRC (NF-SI-0514-10090 and NIHR203331). M.C.P is a Wellcome Trust Senior Fellow in Clinical Science (grant 212252/Z/18/Z). The views expressed are those of the authors and not necessarily those of the NIHR or the Department of Health and Social Care.

## Acknowledgements

We thank the study participants and the clinical research teams who helped with recruitment. We thank Surjeet Singh as the chief study coordinator for TABUL, and Bhaskar Dasgupta as a co-investigator on TABUL who played a leading role in the study recruitment.

