## Supplementary Figures for "Wide-angled proteomics in giant cell arteritis reveals complement pathway activation"

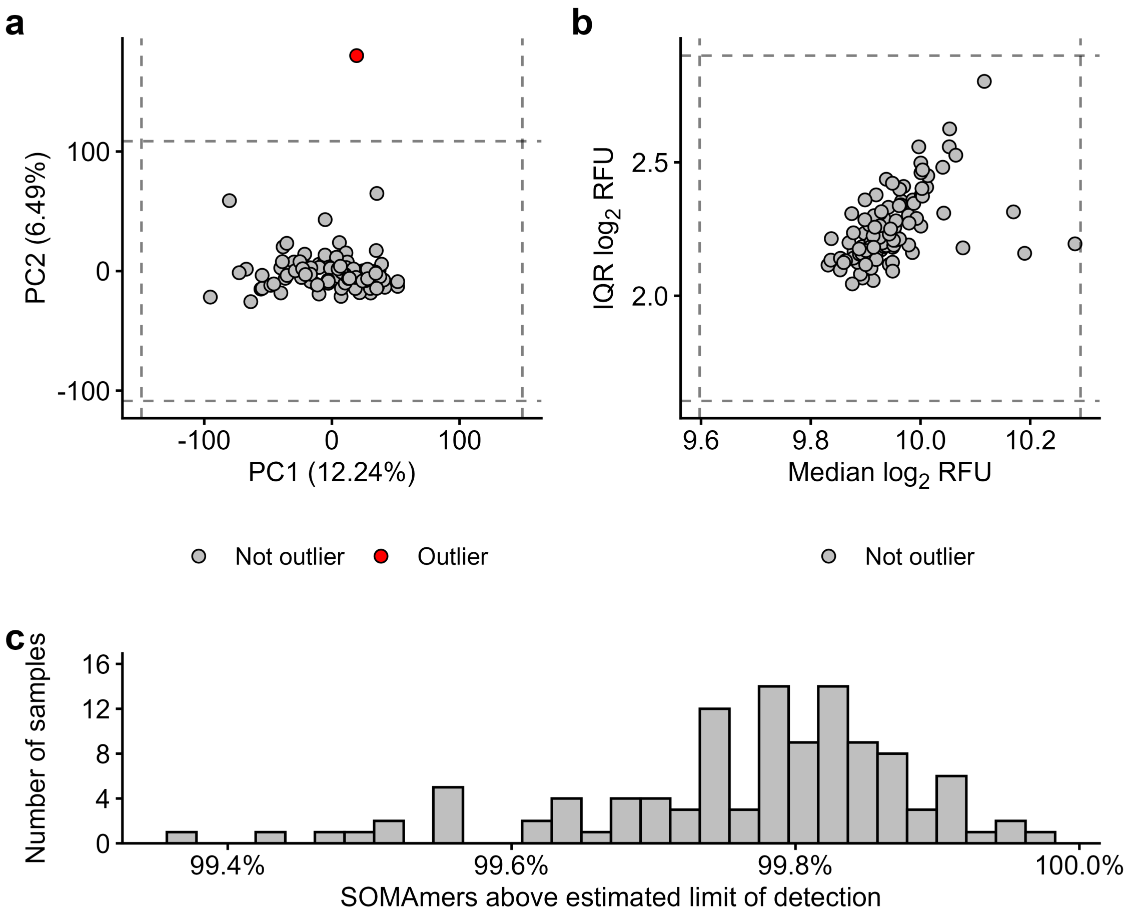


**Supplementary Figure 1. Sample and SOMAmer outlier detection. (a)** Outlier sample identification using the first two proteomic principal components. **(b)** Outlier samples identification using per-sample median and interquartile range protein abundance values. **(c)** Percentage of SOMAmers above their estimated limited of detection (eLOD) in each sample. The dashed lines in (**a**) and (**b**) show +/- 5 standard deviations from the mean value on that axis. Outlier samples are coloured red.

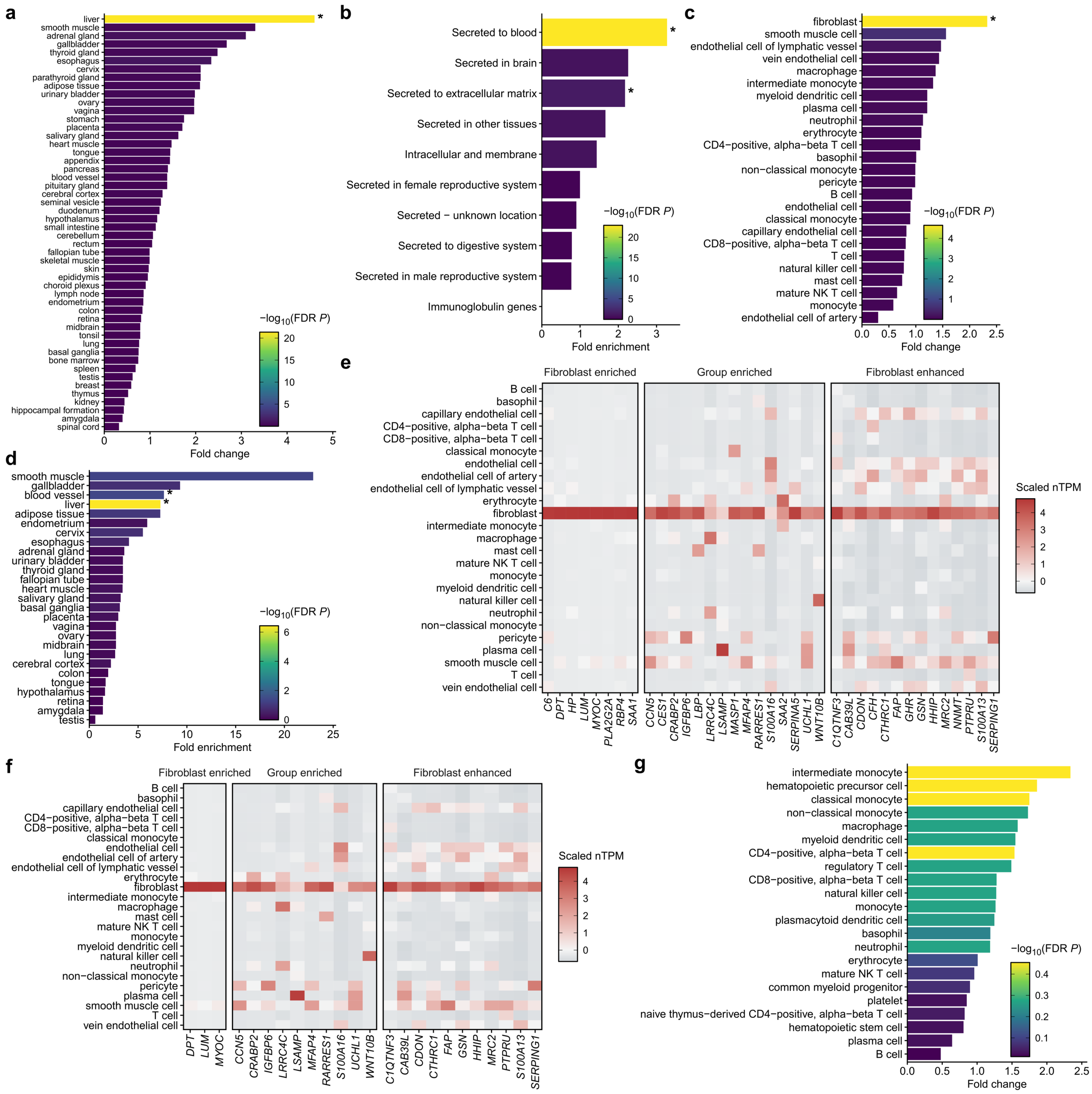


**Supplementary Figure 2. Tissue-, cell- and secretome-specificity of GCA-associated proteins. (a)** Overrepresentation of GCA-associated proteins among tissue-specific genes. Tissue expression based on Human Protein Atlas (HPA) and Genotype-Tissue Expression (GTEx) consensus gene expression references. **(b)** Overrepresentation of GCA-associated proteins among secretome categories. Secretome annotations from the HPA. **(c)** Overrepresentation of GCA-associated proteins among vascular cell types. Vasculature cell-type gene expression reference from Tabula Sapiens v2. **(d)** Overrepresentation of GCA-associated proteins encoded by vascular fibroblast-specific genes among tissue-specific genes. **(e)** Expression of vasculature fibroblast-specific genes in vasculature cell-types. All genes encode GCA-associated proteins. Gene expression from Tabula Sapiens v2. **(f)** Expression of vasculature fibroblast-specific genes in vasculature cell-types, excluding liver-specific genes. **(g)** Overrepresentation of GCA-associated proteins among blood-cell/lymphoid organ specific genes. Cell-type gene expression from Tabula Sapiens v2. * = FDR-adjusted $P < 0.05$ in all panels.

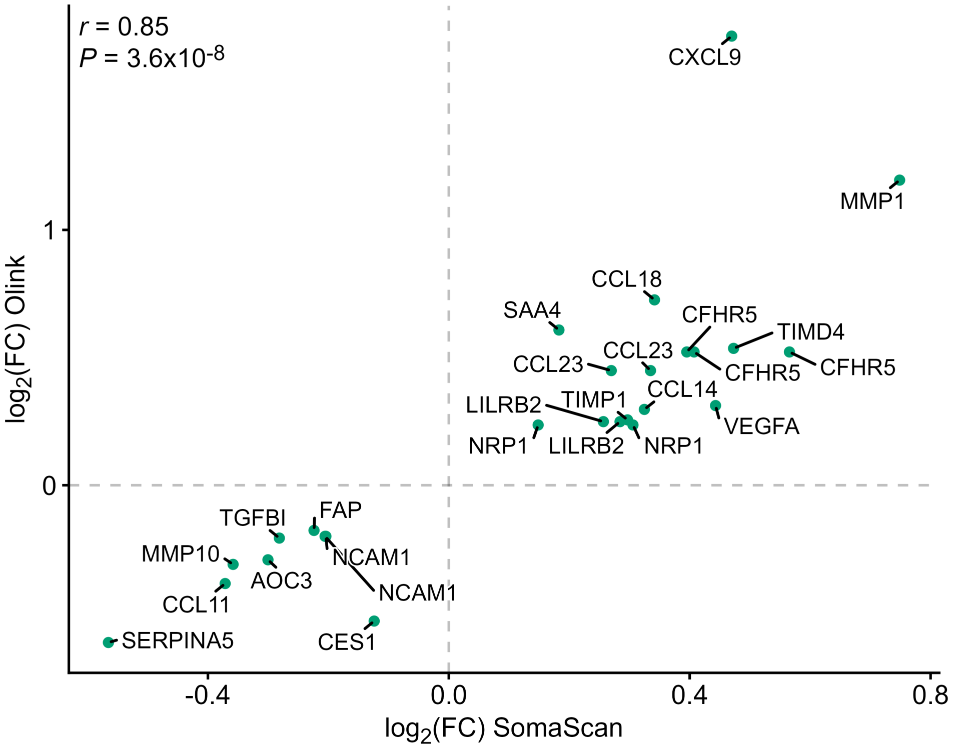


**Supplementary Figure 3. Protein effect sizes in biopsy-confirmed GCA relative to non-GCA TABUL samples measured via antibody- and aptamer-based affinity proteomics.** Shown are targets passing a readjusted multiple testing threshold ($P_{FDR} < 0.05$) based on the union of common proteins between antibody (Olink) and aptamer-based (SomaScan) proteomic platforms. Olink effect sizes taken from Maughan et al (2025). Correlation reported as Pearson's *r* coefficient. Targets labelled according to NCBI gene symbols.

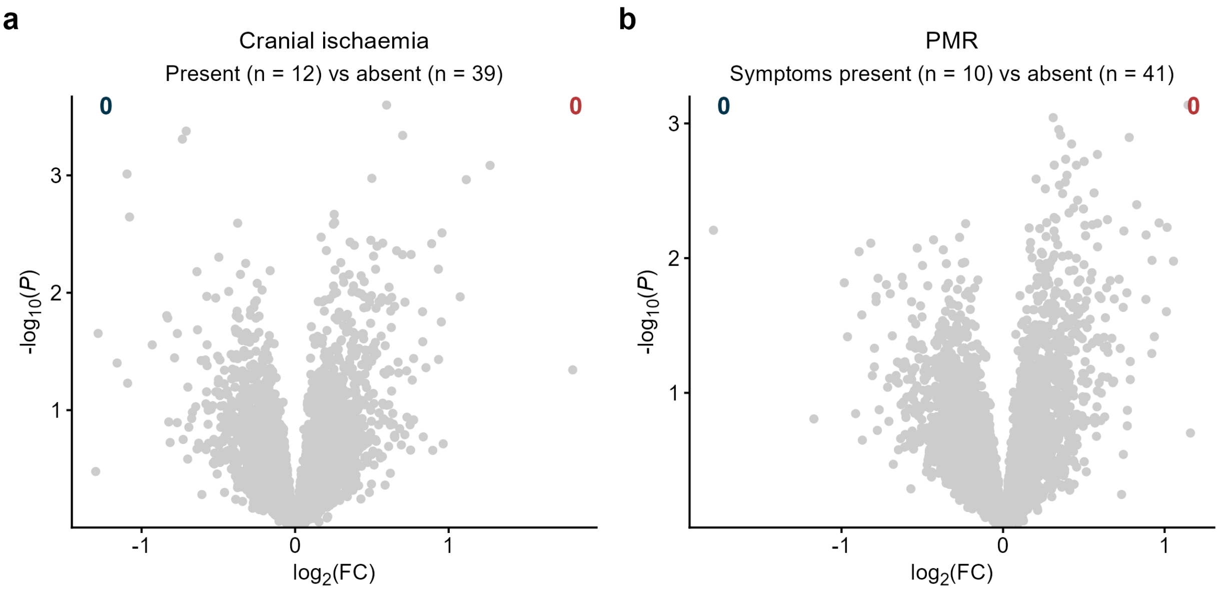


**Supplementary Figure 4. Plasma protein associations with clinical symptoms in GCA patients. (a, b)** Proteomic associations with cranial ischaemia complications (**a**) and the presence of polymyalgia rheumatica symptoms (**b**). No proteins were significantly associated ($P_{FDR} < 0.05$) with either trait.

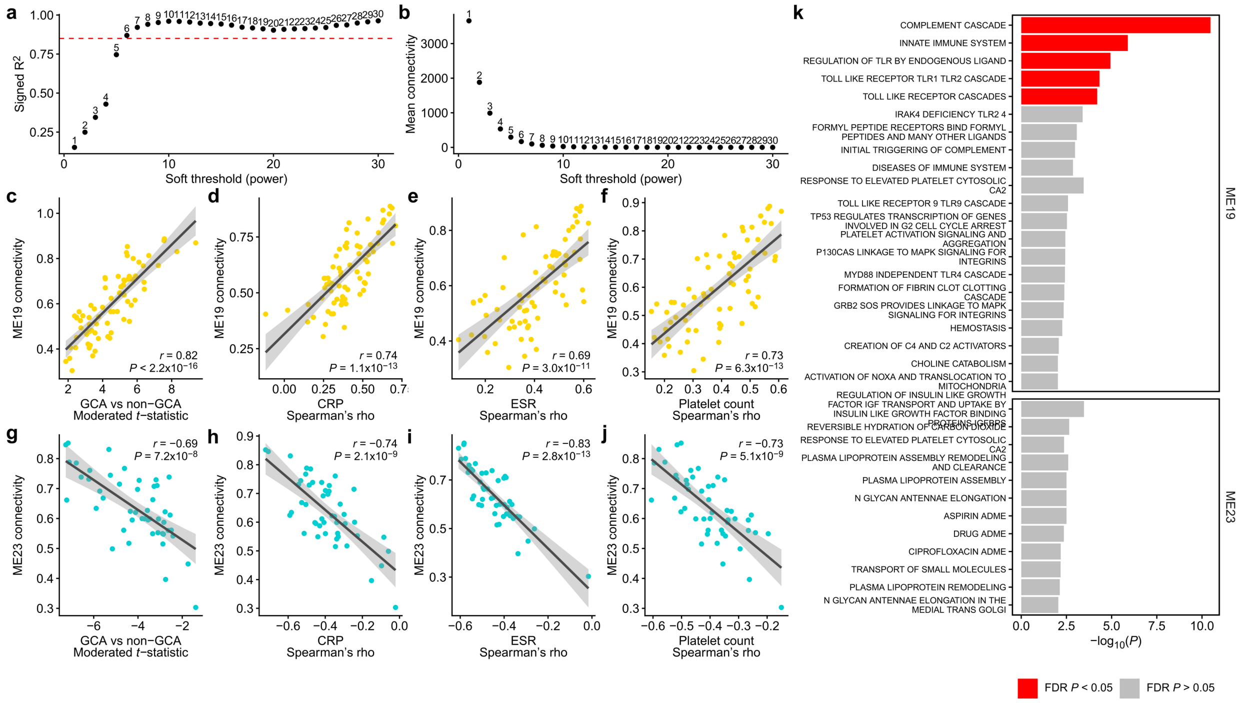


**Supplementary Figure 5. Protein coabundance module analysis. (a)** Scale free topology fit for a range of soft-thresholding powers. Red dashed line shows a signed $R^2$ of 0.85. **(b)** Mean connectivity of potential soft-thresholding powers. **(c-j)** Relationship between module 19 and 23 connectivity and trait associations. Trait associations include diagnosis group (**c, g**), CRP (**d**, **h**), ESR (**e**, **i**) and platelet count (**f**, **j**). CRP = C-reactive protein levels, ESR = Erythrocyte sedimentation rate. Correlations between connectivity and trait associations reported as Pearson's *r* coefficient. **(k)** Overrepresentation of Reactome terms in members of modules 19 and 23. *P* value calculated from a hypergeometric test. *P* values corrected for multiple testing within each module (Benjamini-Hochberg).

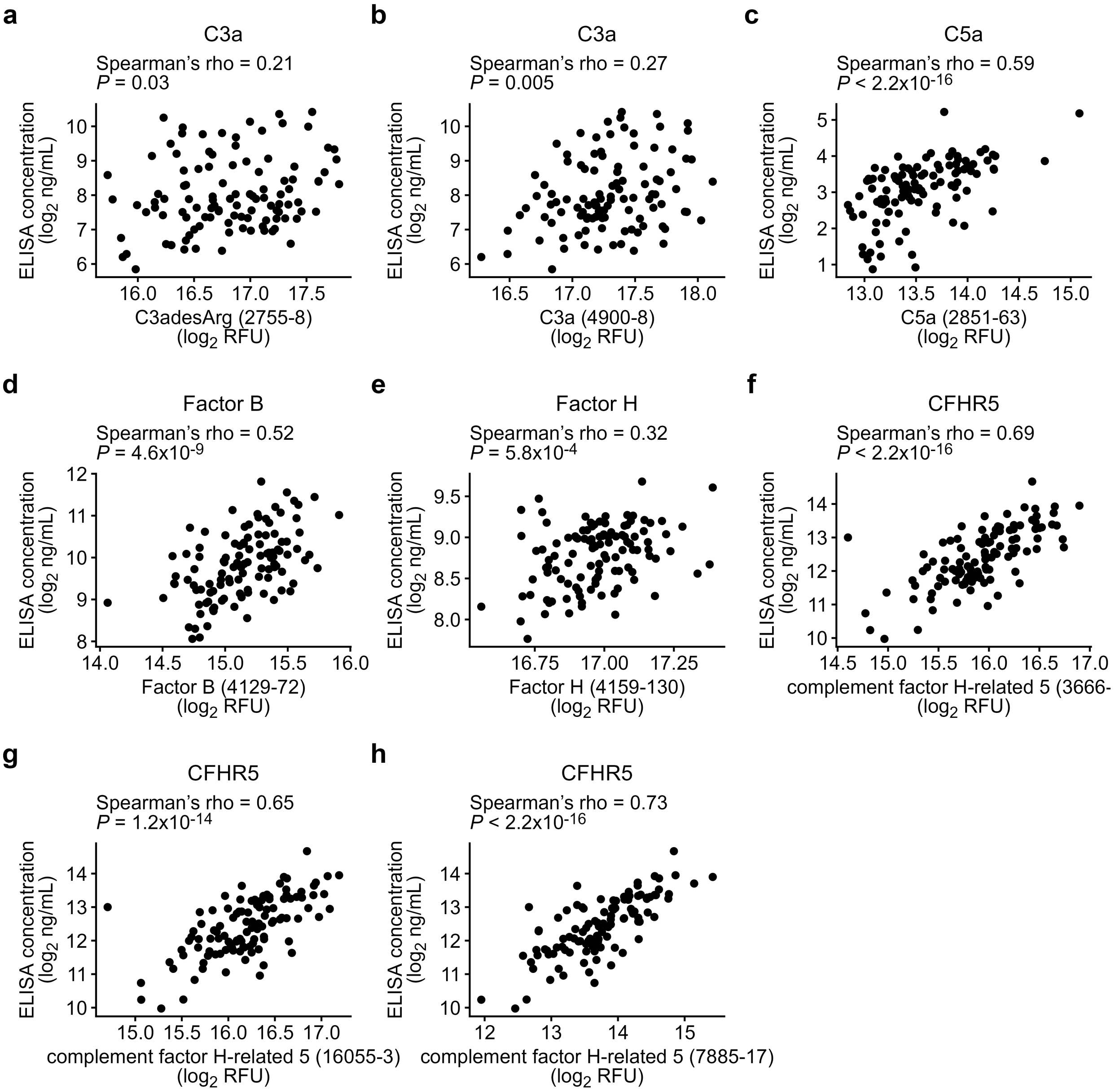


**Supplementary Figure 6. Comparing SOMAmer and ELISA measurements of plasma complement protein levels.** $Log_2$-transformed ELISA concentrations are shown on the y-axes. $Log_2$-transformed relative fluorescent units of the proteins targeted by SOMAmers shown on x-axes. The SOMAmer sequence ID is given in parentheses. Protein measured in corresponding plasma samples from TABUL participants.

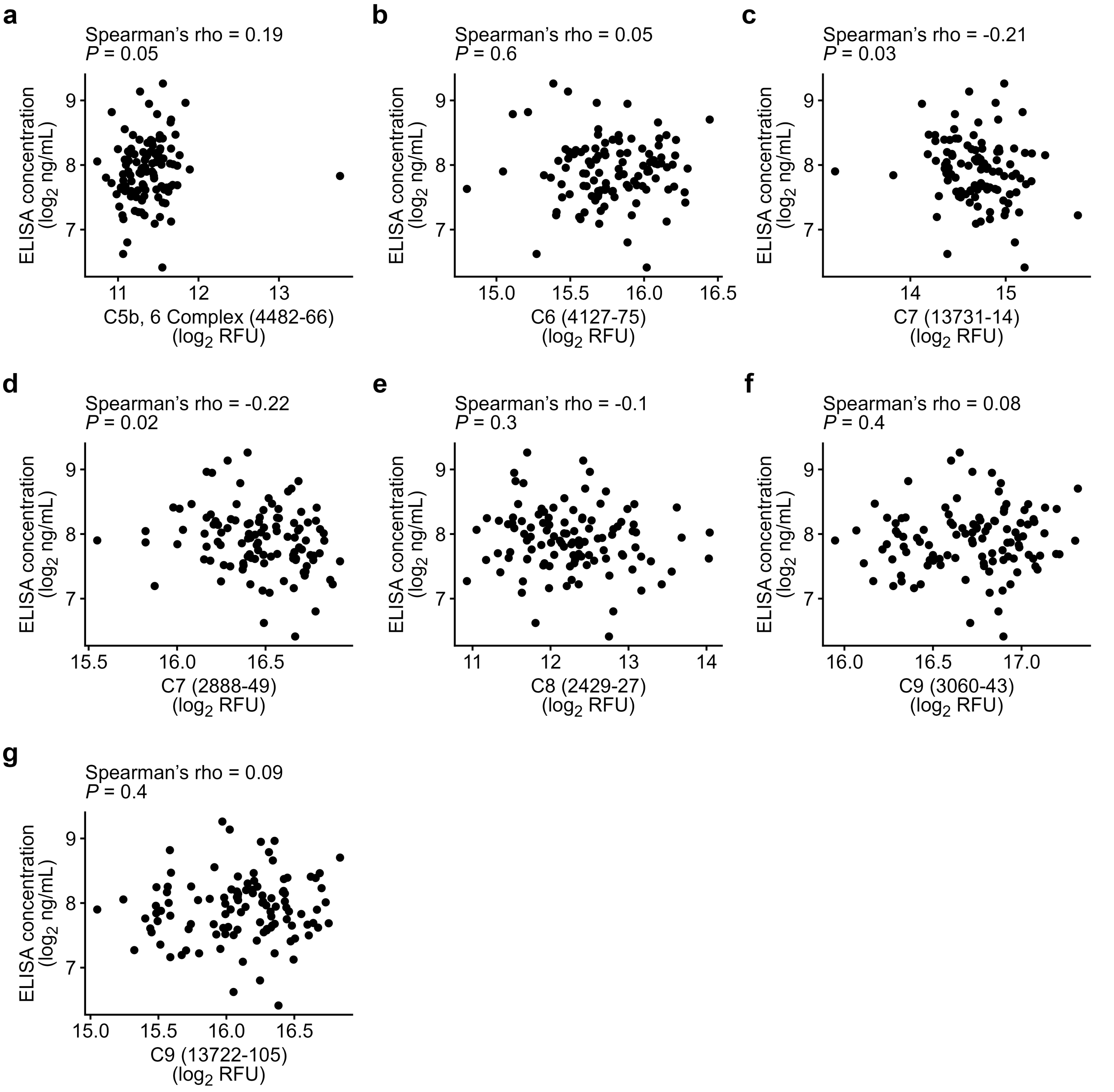


**Supplementary Figure 7. Comparing ELISA measurement of plasma soluble C5b-9 levels with SOMAmer measured terminal complement complex components.** $Log_2$-transformed soluble C5b-9 (sC5b-9) ELISA concentrations are shown on the y-axes. $Log_2$-transformed relative fluorescent units of terminal complement complex components targeted by SOMAmers shown on x-axes. The SOMAmer sequence ID is given in parentheses. Protein measured in corresponding plasma samples from TABUL participants.

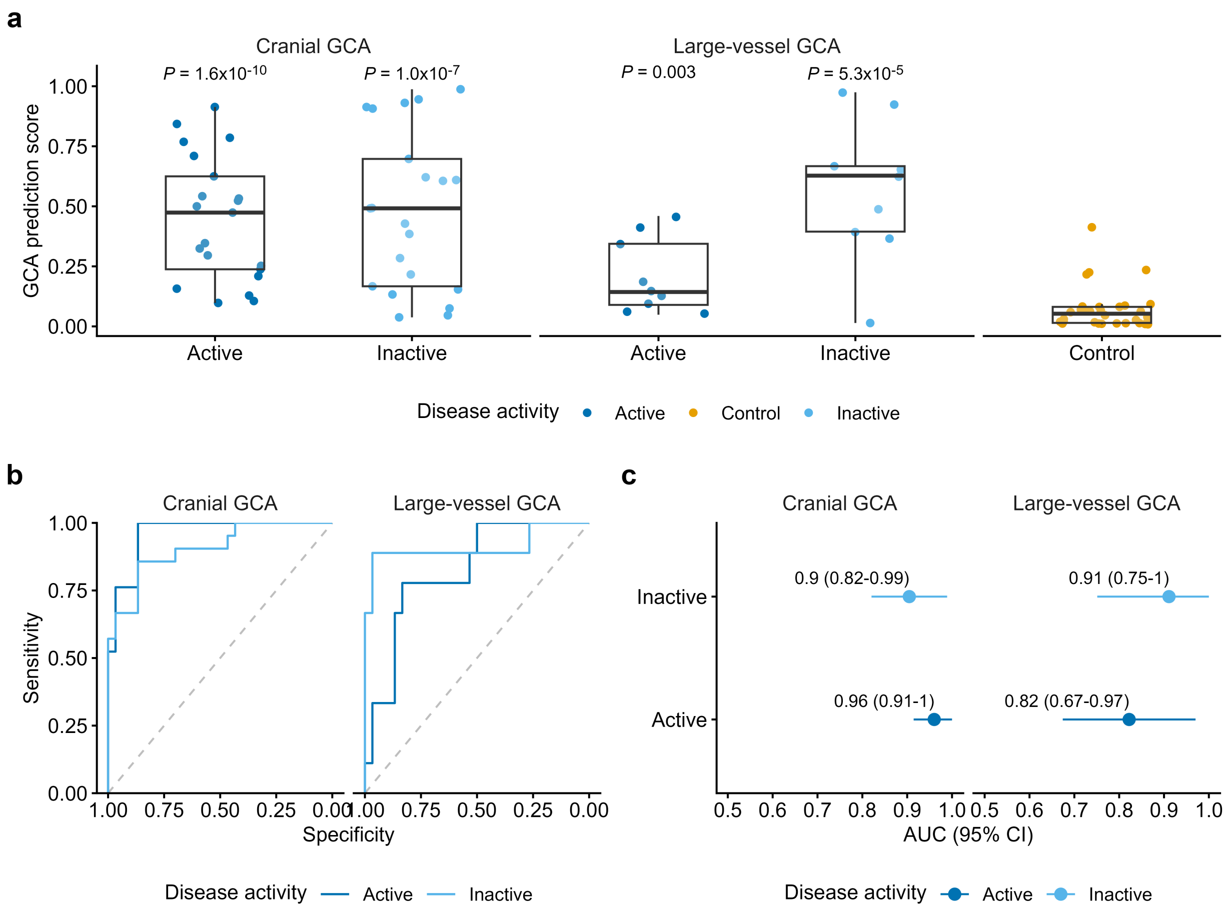


**Supplementary Figure 8. Model predictions on external cranial- and large-vessel GCA samples. (a)** Model predictions on external GCA proteomic data generated by Cunningham et al (2024) stratified into cranial GCA and large-vessel GCA. *P* values calculated from a two-sided Wilcoxon rank-sum tests comparing Control sample predictions to active/inactive cranial- and large-vessel GCA samples. *P* values Bonferroni corrected by four tests. **(b)** ROC curves for model predictions distinguishing active and inactive cranial- and large-vessel GCA from Control samples. **(c)** AUCs for thresholds shown in (**b**).

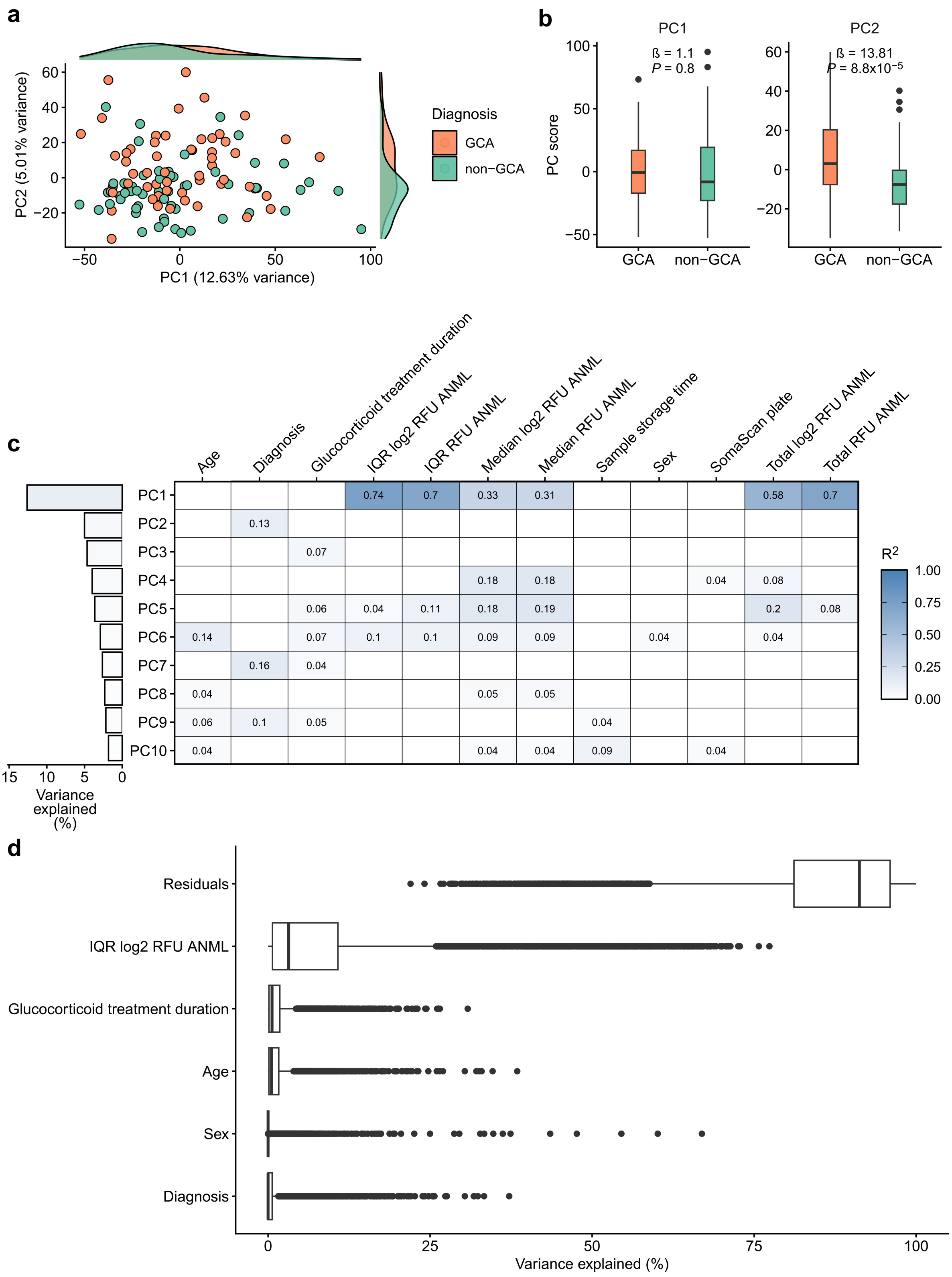


**Supplementary Figure 9. Explainable sources of plasma protein abundance variation. (a)** First two protein abundance principal components (PCs). Each point shows a sample labelled by GCA diagnosis. **(b)** First two PC scores stratified by GCA diagnosis. Association measured using linear regression. **(c)** Univariate linear regression of measurable sources of variation against the first ten protein abundance PCs. The top panel shows the variance explained by each consecutive PC. $R^2$ values are shown for significant associations ($P < 0.05$). **(d)** Explainable protein abundance variation. Each point shows an individual SOMAmer.
